# Genetic association and machine learning improves discovery and prediction of type 1 diabetes

**DOI:** 10.1101/2024.07.31.24311310

**Authors:** Carolyn McGrail, Timothy J. Sears, Parul Kudtarkar, Hannah Carter, Kyle Gaulton

**Author notes:** Corresponding authors: Contact: Carolyn McGrail,; Kyle J Gaulton. Authors contributed equally to this work (Carolyn McGrail, Timothy J. Sears). Authors jointly supervised this work (Hannah Carter, Kyle Gaulton).

## Abstract

Type 1 diabetes (T1D) has a large genetic component, and expanded genetic studies of T1D can lead to novel biological and therapeutic discovery and improved risk prediction. In this study, we performed genetic association and fine-mapping analyses in 817,718 European ancestry samples genome-wide and 29,746 samples at the MHC locus, which identified 165 independent risk signals for T1D of which 19 were novel. We used risk variants to train a machine learning model (named T1GRS) to predict T1D, which highly differentiated T1D from non-disease and type 2 diabetes (T2D) in Europeans as well as African Americans at or beyond the level of current standards. We identified extensive non-linear interactions between risk loci in T1GRS, for example between HLA-DQB1*57 and *INS,* coding and non-coding HLA alleles, and *DEXI, INS* and other beta cell loci, that provided mechanistic insight and improved risk prediction. T1D individuals formed distinct clusters based on genetic features from T1GRS which had significant differences in age of onset, HbA1c, and renal disease severity. Finally, we provided T1GRS in formats to enhance accessibility of risk prediction to any user and computing environment. Overall, the improved genetic discovery and prediction of T1D will have wide clinical, therapeutic, and research applications.

---

Type 1 diabetes (T1D) is an autoimmune disease characterized by immune infiltration of the pancreatic islets and beta cell destruction, leading to lifelong dependance on insulin therapy^1^. Onset of T1D is typically during early life or adolescence, but can also occur in adults^2^. While both genetic and environmental factors contribute to the development of T1D, the underlying etiology is not fully understood^1^. Determining individuals at risk for T1D can help prevent diabetic ketoacidosis at onset, which can lead to life-long complications, as well as to inform selection for preventative therapies such as teplizumab^3–5^. Typically T1D is accompanied by autoantibodies (Aab) against islet-specific proteins which can be detected in blood prior to disease onset and used as biomarkers^6^. However, islet Aab can be transient, are less frequently found in adult-onset cases, and are not fully predictive of T1D^6,7^. Furthermore, the detection of islet Aab is limited until after T1D progression has started where many individuals already have reduced beta cell function, and therefore earlier prediction of disease is needed.

Inherited genetic risk factors can also predict the development of T1D. Genetic variants in class I and II MHC genes are the largest risk factors for T1D^8,9^, most notably the HLA-DRB1*0301-DQB1*0201 (DR3-DQ2) and HLA-DRB1*0401-DQB1*0302 (DR4-DQ8) haplotypes which when inherited together increase risk of T1D over 16-fold^8^. Outside of the MHC locus, T1D is highly polygenic with over 90 genomic loci such as *INS* and *PTPN22* harboring variants affecting T1D risk^10–13^. The heritability of T1D, however, remains incompletely described^14^, and therefore additional T1D-associated variants likely remain to be discovered. In addition, risk loci detected through association studies contain many associated variants due to linkage disequilibrium and at best indicate the presence of one or more unknown causal variants^15^. Larger genetic association studies can boost statistical power to detect new risk loci while detailed fine-mapping analyses can narrow down candidate causal variants for these signals, which together can enhance the ability to predict T1D using genetic variation^15^.

Genetic variants associated with T1D risk can be used to construct genetic risk scores (GRS)^16–18^, which are summarizations of the T1D genetic risk of an individual used to predict the development of T1D. The utility of a GRS as a diagnostic tool can, for example, aid in selection for preventative therapies, complement interpretation of blood biomarkers, and distinguish between forms of diabetes^16,18,19^. Previous studies have created risk scores for T1D which accurately discriminate T1D from non-diabetes, as well as T1D from T2D^16,18,20^. Improvements to existing T1D GRS, however, can further enhance the utility and accessibility of GRS. Current T1D GRS are calculated using the additive sum of effects for risk alleles of an individual and, outside of several known interactions in specific class II MHC alleles such as DR3-DQ2 and DR4-DQ8, do not consider non-linear interactions between variant alleles^16,18,20^. Furthermore, existing T1D GRS are not straightforward to calculate from imputed genotypes derived from reference panels such as TOPMed and require extensive phasing of HLA haplotypes and use of proxy SNPs^18,20^. These complexities can further lead to inconsistencies resulting in ambiguity of the HLA alleles in an individual^18^. These issues limit the utility of current T1D GRS in both basic research and clinical settings.

In this study, we performed the most comprehensive genetic association study of T1D to date using genome-wide association and fine-mapping analyses in 817,718 samples as well as detailed fine-mapping of the MHC locus in 29,746 samples, which revealed 199 total T1D risk variants. We then utilized T1D risk variants to train a machine learning (ML) model, named T1GRS, to predict T1D. The T1GRS model exceeds diagnostic ability compared to the current standard T1D GRS2 and reveals novel interactions both at the MHC locus and other loci genome wide. T1GRS also improves accessibility of T1D prediction by using publicly available imputation panels and by providing containerized pipelines. Finally, we use predictions from T1GRS to define genetic sub-groups of T1D individuals which have distinct clinical features.

## Results

### Genome-wide association and fine-mapping identifies novel T1D risk loci and signals

We performed a T1D association study of 817,718 samples (20,355 T1D cases and 797,363 non-diabetes) using a meta-analysis of 9 cohorts consisting of individuals of European ancestry (**Supplementary Table 1**). Variants were imputed into the TOPMed v3 reference panel or, for UK Biobank, the HRC reference panel, and up to 62.1M variants with minor allele frequency (MAF)>1×10^−5^ were considered in association tests. In total, we identified 92 loci that reached genome wide significance (P<5×10^−8^) of which 79 were previously known T1D risk loci and the other 13 had not, to our knowledge, been previously reported (**Fig 1A,B**).

**Figure 1.**
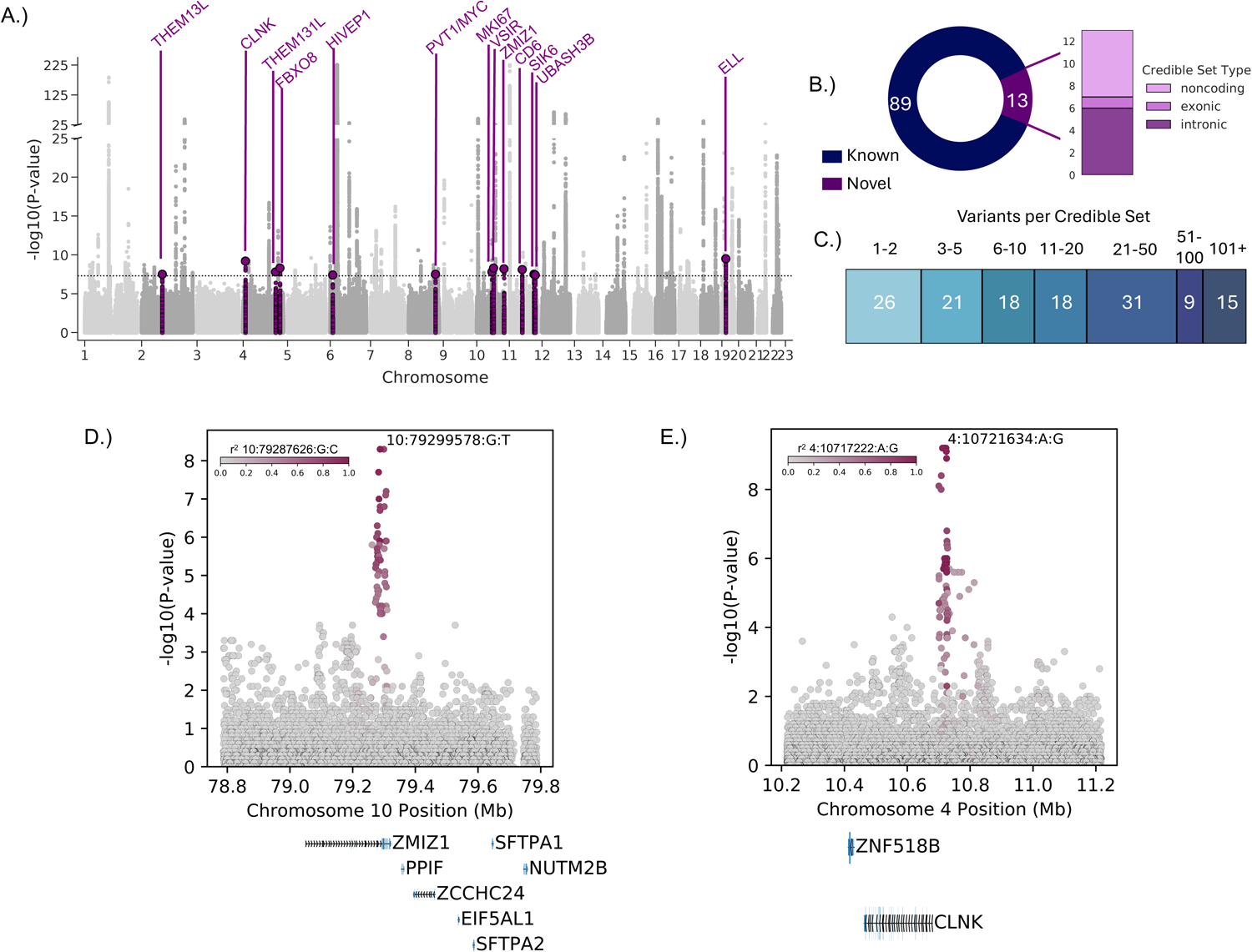
Genome-wide association and fine-mapping of type 1 diabetes. **A.** Genome-wide association analysis of type 1 diabetes (T1D). *P* values are from the marginal T1D association statistics in a meta-analysis of *n* = 817,718 samples. Novel loci with p-values less than 5×10^−8^ are colored purple and labeled. Variants with p-value less than 1×10^−225^ were capped at this threshold for display. **B.** In total there are 102 T1D loci including 89 previously established loci and 13 novel loci. The genomic annotation category of the novel loci is shown to the right. **C.** Number of credible set variants for the 138 signals identified after fine-mapping 102 T1D loci with SuSIE. **D,E.** Locus plots showing T1D association for the novel loci *ZMIZ1* and *CLNK*. The color of each variant represents the linkage disequilibrium (r^2^) with the lead variant at the locus.

We next performed fine-mapping of likely causal variants at the 92 identified loci, plus an additional 10 previously established T1D loci that did not reach genome-wide significance in our study, using SuSIE (**see methods**). Across all 102 loci, we identified 138 independent risk signals, where several loci such as *IL2RA, INS, PTPN22,* and *IFIH1* had more than 2 independent signals (**Supplementary Table 2**). At each independent signal, we derived ‘credible sets’ of variants likely causal for the signal (**Supplementary Table 3**). Over half (79/138) of the credible sets had 15 or fewer variants and over a third (47/138) had 5 or fewer variants (**Fig.1C**). Signals with larger credible sets (>125 variants) included many of the loci that did not reach genome-wide significance in our study and likely have lower fine-mapping resolution due to weaker association. Overall, there was a high degree of concordance with previous fine-mapping, where ∼97% of independent signals at known loci in this study overlapped signals in a previous genome-wide association study^10^. At several signals the fine-mapping resolution was improved, likely due to both the larger sample size as well as the method used in fine-mapping. For example, at the *CTRB1/2* locus, fine-mapping resolved a single causal variant (rs55993634, PIP=.989), whereas this same variant previously had lower probability^10^. At the *INS* locus, fine-mapping resolved likely causal variants at all three independent risk signals (rs689 PIP=.87; rs7948458 PIP=.96; rs11042966 PIP=.99). At a small number of loci there was limited overlap in credible sets compared to previous studies, notably the *UBASH3A* locus, where reported signals have complex patterns of linkage disequilibrium (LD)^10^.

We annotated credible set variants at the 13 novel loci to understand potential molecular mechanisms of these loci. At seven loci a candidate variant mapped within a protein-coding gene and the remaining six mapped to intergenic sequence. At the *ZMIZ1* locus, candidate variants mapped in the intron of the *ZMIZ1* gene which is also involved in T2D risk and affects beta cell function and glucose homeostasis (**Fig 1D**)^21^. At another locus, candidate variants mapped to an intron of a long noncoding RNA *PVT1* downstream from *MYC* that has been shown to be associated with renal diabetic complications^22^. Loci that were intergenic included variants near *CLNK* (**Fig 1E**), a mast cell immunoreceptor signal transducer that plays a role in the regulation of immunoreceptor signaling. Credible set variants at this locus further overlapped accessible chromatin sites in pancreatic and immune cell types (**Supplementary Table 4**).

In total these results provide expanded genetic association and fine-mapping of T1D risk and revealed novel risk loci and independent risk signals at both known and novel loci.

### Fine-mapping of the MHC locus identifies additional class I and II risk signals

Given the particular importance of the MHC locus to T1D genetic risk, we next performed detailed fine-mapping to identify additional T1D risk signals at this locus. We utilized genotypes from 29,746 T1D case and control individuals of European ancestry from five publicly available cohorts (**see Methods)**. After quality control, we imputed variant genotypes into the HLA reference panel in the Michigan Imputation Server containing 55,614 variants including two- and four-digit HLA alleles, amino acid residues, and intragenic variants^23^.

Given the extreme associations and extensive LD in the MHC region, methods such as SuSIE are currently not as amenable to fine-mapping this locus. We therefore instead performed a stepwise conditional analysis by iteratively adding the most significant variant into the model and re-performing association tests (**see Methods**). In total, this process revealed 23 independent signals associated with T1D at genome-wide significance (p<5×10^−8^) (**Fig. 2A,B, Supplementary Table 5,6**). For each signal we then derived ‘credible sets’ of variants likely causal for the signal using a Bayesian approach. As expected, these signals primarily consisted of variants in LD with class I and class II MHC alleles that have established roles in T1D (**Fig. 2C, Supplementary Table 5).** For example, signals represented the amino acid change in *HLA-DQB1* at position 57 (rs1064173 r^2^=0.65)^24^ and *HLA-DRB1* at position 13 (rs1391373 r^2^=0.763)^25^, and known T1D risk and protective alleles including *HLA-DQB1*\*02:01 (rs9273530 r^2^=0.991), *HLA-DQB1*\*06:02 (rs9273375 r^2^=0.801), *HLA-A*\*24:02 (rs17185657 r^2^=0.973)^8^, and *HLA-B*\*39. We also identified a small number of signals not linked to any known T1D risk or protective MHC alleles, including a non-coding signal indexed by rs9276235 that maps to an intergenic region in between *HLA-DRB1* and *HLA-DQA2* (**Supplementary Fig. 1A**).

**Figure 2.**
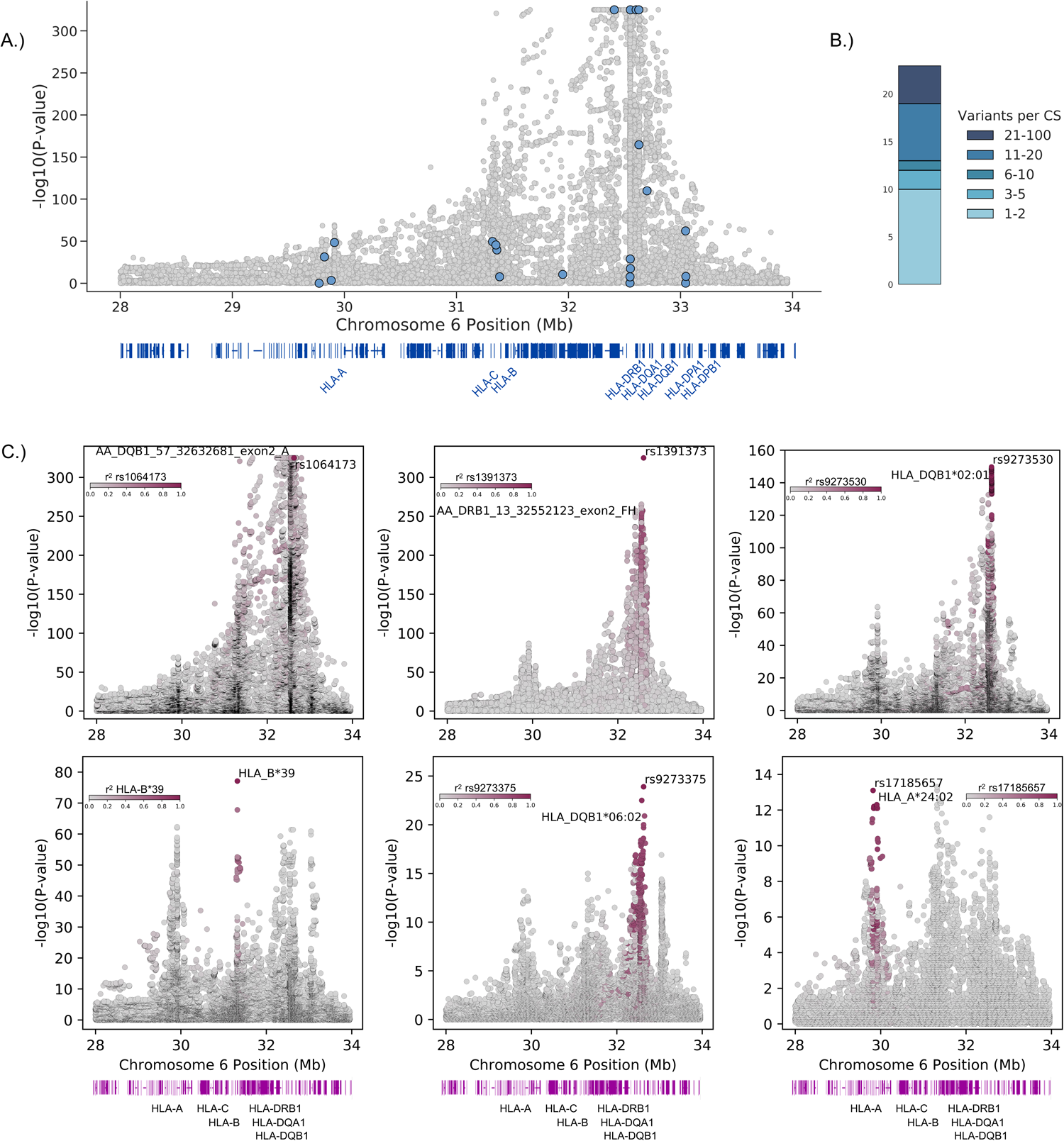
Genetic association of T1D at the MHC locus. **A.** T1D association at the MHC locus. P values are from the marginal T1D association statistics frm meta-analysis of *n* = 29,746 samples. The lead variant for each signal is colored blue and signals with a p-value less than 1×10^−325^ were capped at this threshold for display. The gene map shows the location of class I and II HLA genes. **B.** The number of variants per credible set are shown for the 23 signals identified in conditional fine-mapping. **C.** Locus plots with conditional T1D association for signals linked to known T1D HLA or amino acid associations. Locus plots are colored by linkage (*r^2^*) to the lead variant for each signal. The gene maps show the location of class I and II HLA genes.

As novel signals at the MHC locus may be obscured by partial LD with known risk due to the large and extensive associations in the region, we next performed a second round of stepwise analyses at the HLA locus after first pre-conditioning on 70 known class I and class II MHC risk alleles and 20 known DR3/DR4 haplotypic and allelic interactions (**See methods, Supplementary Table 7-9**). These analyses revealed four additional novel signals (p<5×10^−8^) not linked to any known HLA risk alleles (**Supplementary Fig. 1B**), one of which is a coding signal consisting of the amino acid residue 71 in *HLA-DRB1*^26^. The other three novel signals map fully to non-coding sequence, and we annotated credible set variants for these signals using published maps of accessible chromatin sites and transcription factor (TF) binding. For example, at the novel signal near *HLA-A,* credible set variant rs7763052 (PIP=0.173) overlaps an accessible chromatin site active in effector CD8^+^ T cells and NK cells (**Supplementary Fig. 1C**)^27^. At another novel signal located upstream of *HLA-DRB1* candidate variant rs9270965 (PIP=0.987) preferentially bound to SOX3 in a published high-throughput SELEX-seq assay^28^. Altogether, the use of an HLA-specific imputation panel resolved independent signals at the HLA locus including several coding and non-coding signals not previously implicated in T1D risk.

### A machine learning model improves classification of T1D from non-diabetes and T2D

We next sought to use known and novel risk alleles at the MHC locus and genome-wide to predict T1D. We developed a machine learning model using gradient boosting to predict T1D, which we named T1GRS. The T1GRS model comprises a total of 199 T1D-associated variants, including 70 known HLA risk variants, 27 additional HLA variants identified in conditional analyses, and lead variants at the primary signals of 102 risk loci genome-wide (**see Methods, Supplementary Table 10**). We trained both a “full” model using all 199 variants in addition to sex, population structure captured by the first four principal components derived from genotyping data, and cohort label, as well as a “reduced” model using just the 199 variants for application in settings where the other variables are not available or relevant. In addition to a model consisting of all 199 variants, we also trained sub-models consisting of only (i) the 97 HLA variants or (iii) the 102 variants mapping outside the HLA locus (non-MHC). We evaluated the ability of each model to differentiate T1D from non-diabetes in 29,746 individuals (10,107 T1D and 19,639 non-diabetes) using 10-fold cross-validation.

We evaluated T1GRS for the ability to discriminate T1D from non-diabetes using 29,746 European-ancestry samples from five cohorts where T1D cases had a range of age of diagnosis (**Supplementary Table 1**). When testing the ability of T1GRS to differentiate T1D from non-diabetes, there was overall strong discrimination when using all 199 variants (full model area under the curve AUC=0.931) (**Fig. 3A**). When calculating precision-recall in T1GRS, there was an Average Precision (AP) score of 0.867 (**Supplementary Fig. 2A**). We observed similar discrimination of T1D when using the 97 MHC variant model (AUC=0.915, AP=0.835) (**Fig. 3B, Supplementary Fig. 2B**). Surprisingly, we also observed reasonable discrimination of T1D when using only the 102 variants outside of the HLA locus, although the average precision was lower (AUC=0.800, AP=0.660) (**Fig. 3C, Supplementary Fig. 2C**).

**Figure 3.**
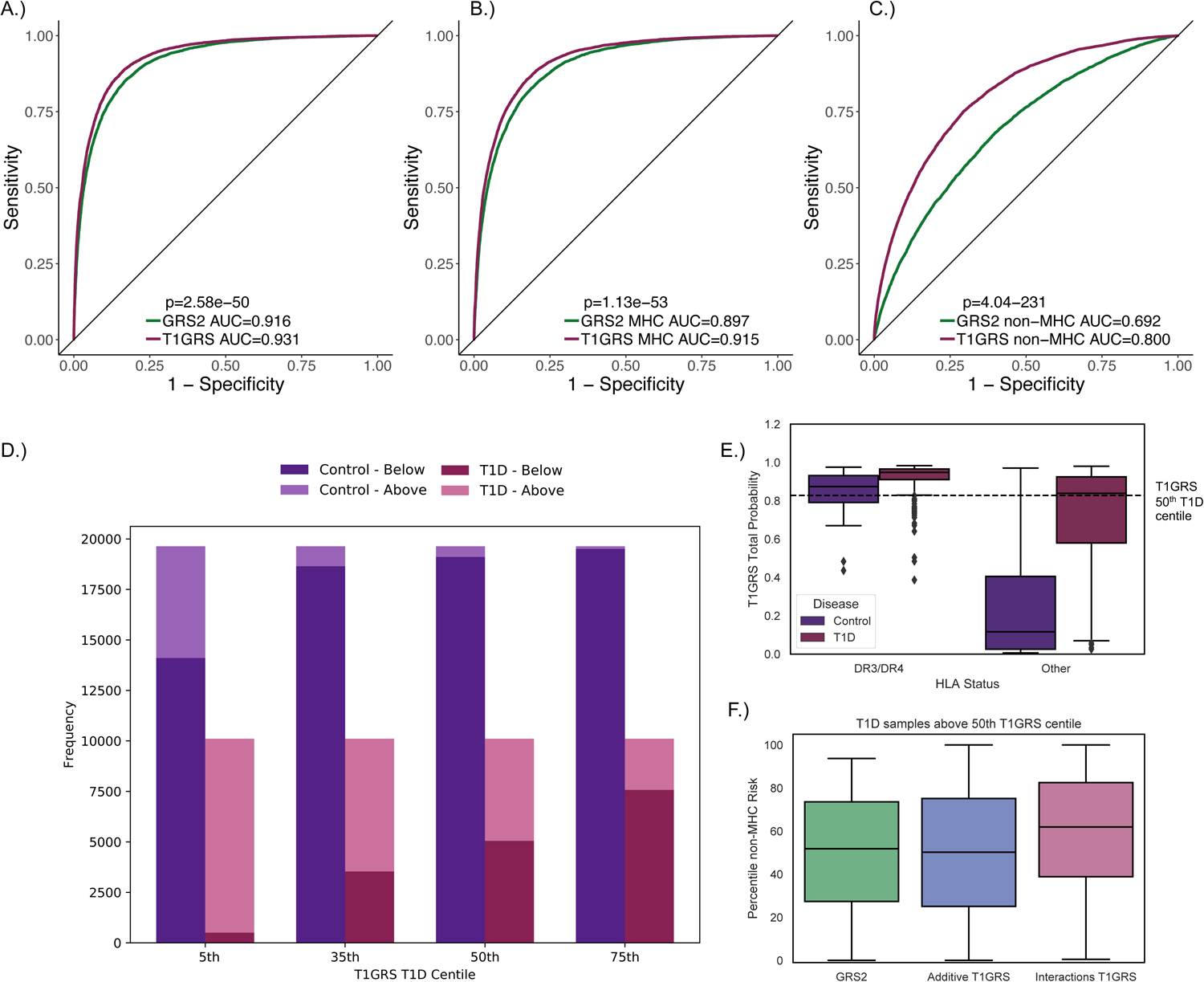
Genetic prediction of T1D using T1GRS. Receiver operating characteristic (ROC) curves assessing the accuracy of predicting T1D from non-diabetes using T1GRS and T1D GRS2. The AUC for T1GRS is colored purple while the existing T1D GRS2 is colored green. P values comparing predictive ability of GRSs are calculated using the de-Long test. The AUCs for T1GRS and GRS2 are shown for **A.** All variants, **B.** MHC-only variants, and **C.** non-MHC genome-wide variants. **D.** Percentile thresholds for T1GRS probability calculated in individuals with T1D compared with non-diabetes. **E.** T1GRS probability in individuals with HLA-DQ proxy SNP ambiguity in T1D GRS2 separated by high-risk HLA-DR3/DR4 (left) and all other HLA alleles (right). **F.** Non-MHC percentile risk in individuals with T1D above the T1GRS total 50^th^ T1D threshold. Green represents GRS2 non-MHC percentile risk, blue is the percentile scaled to an additive effect of T1GRS non-MHC variants, and pink is the non-MHC T1GRS probability percentile that includes non-linear interactions. GRS2 scores for individuals who fall above the 50^th^ T1D percentile in T1GRS.

We next compared the predictive ability of T1GRS to the previously reported T1D risk score GRS2^18^. There was significantly improved ability of the T1GRS model to discriminate T1D from non-disease compared to GRS2 when using all variants (GRS2 Total AUC= 0.916, p=2.58×10^−50^) and MHC variants (GRS2 MHC AUC= 0.897, p=1.13×10^−53^) (**Fig. 3A-B**). The greatest improvement, however, was observed when considering only non-MHC variants genome-wide, where T1GRS dramatically improved over GRS2 (GRS2 non-MHC AUC=0.692, p=4.04×10^−231^) (**Fig. 3C**). Similarly, the average precision across each model was higher in T1GRS compared to GRS2, which was again most prominent in the non-MHC model (GRS2 all variant AP=0.839, MHC AP=0.803, non-MHC AP=0.517) (**Supplementary Fig. 2A-C**).

We then examined the application of the “reduced” T1GRS model to predict T1D. In the training data, this T1GRS model significantly outperformed GRS2 using all variants (AUC=0.919, p=1.46×10^−4^) and non-MHC variants (AUC=0.714, p=2.75×10^−20^), although had slightly reduced performance among MHC-only variants (AUC=0.894, p=7.84×10^−3^) **(Supplementary Fig. 3A-C)**. We next applied this T1GRS model to an independent set of 263 T1D case and non-diabetic samples of European ancestry from the nPOD biorepository^29^, and again T1GRS strongly differentiated T1D from non-disease (AUC=0.887, MHC AUC=0.874, non-MHC AUC=0.727) **(Supplementary Fig. 3D-F)**. The T1GRS model outperformed GRS2 in the independent samples in all comparisons, particularly in the non-MHC model (GRS2 Total AUC=0.882; GRS2 MHC AUC=0.870; GRS2 non-MHC AUC=0.678). **(Supplementary Fig. 3D-F)**.

As genetic risk scores have been used to help distinguish type 1 and type 2 diabetes (T2D) to avoid misdiagnosis^18,20^, we next evaluated the ability of T1GRS to differentiate T1D from T2D cases. For these comparisons we used 1,999 T2D samples in the WTCCC1 cohort and the 115 T1D samples in the nPOD biorepository described above (**Supplementary Fig. 3G-I**)^30^. The T1GRS model accurately differentiated T1D and T2D using all variants (AUC=0.876) (**Supplementary Fig. 3G**). As with the non-diabetes comparisons, this predictive ability was largely driven by MHC variants (AUC=0.865) (**Supplementary Fig. 3H**) and to a lesser extent the non-MHC variants (AUC=0.661) (**Supplementary Fig. 3I**). The ability of T1GRS to discriminate T1D from T2D was similar to T1D GRS2 across all comparisons (GRS2 AUC=0.867, MHC AUC=0.867; non-MHC AUC=0.605) **(Supplementary Fig. 3G-I)**.

Finally, we evaluated the cross-ancestry portability of T1GRS by measuring its ability to distinguish T1D from non-diabetes in African American individuals. It has been previously argued that European GWAS does not effectively capture T1D risk in African Americans^31^. We tested T1GRS on 284 T1D and 404 non-diabetes individuals of African American ancestry and, not surprisingly, T1GRS discriminated T1D in this population to a significantly lower degree compared to Europeans (AUC=0.844, AUC=0.931, p=1.45×10^−08^) **(Supplementary Fig. 3J)**. The ability of T1GRS to predict T1D in African Americans was almost solely driven by MHC variants (AUC=0.833), as the non-MHC variants had very limited predictive ability (AUC=0.583). However, the predictive ability of T1GRS in African Americans was the same as a published T1D risk score derived from a T1D GWAS performed in African American individuals (AUC=0.846, p=0.879) **(Supplementary Fig. 3K)**, suggesting that T1GRS can be used to predict T1D in African Americans at the same level as current standards^31^.

In total, this reveals that T1GRS can discriminate T1D from T2D or non-diabetes at the same or greater level than the current standard GRS in European and African American ancestry individuals including, for Europeans, a marked improvement in predictions when using genetic risk outside of the MHC locus.

### Application of T1GRS as a diagnostic test for T1D in European individuals

We next determined the diagnostic ability, specificity, and sensitivity of T1GRS using the 29,746 T1D and non-disease individuals (**Table 1**). The output from the T1GRS model ranges from 0-1 and describes the probability that the individual will have T1D. When comparing the ability of the T1GRS model to discriminate T1D from non-diabetes, at a probability threshold of 0.828 there was 50% sensitivity and 97.31% specificity for T1D. The Youden index is a formula that can indicate diagnostic feasibility of a GRS when greater than 0.50. The maximum Youden index for T1GRS was 0.723 at a probability threshold of 0.248, corresponding to 89% sensitivity and 83% specificity for T1D, which is an improvement over the maximum Youden Index for GRS2 of 0.683.

**Table 1.** T1D prediction using T1GRS.

| **T1GRS probability** | **T1D Centile** | **Non-Disease Centile** | **Sensitivity (%)** | **Specificity (%)** | **Youden Index** |
| --- | --- | --- | --- | --- | --- |
| 0.116742 | 5 | 71.83 | 95.00 | 71.83 | 0.6683 |
| 0.229681 | 10 | 81.85 | 90.00 | 81.85 | 0.7184 |
| 0.347085 | 15 | 87.00 | 85.00 | 87.00 | 0.7200 |
| 0.450841 | 20 | 89.98 | 80.00 | 89.98 | 0.6998 |
| 0.54173 | 25 | 92.14 | 75.00 | 92.14 | 0.6714 |
| 0.617035 | 30 | 93.69 | 70.00 | 93.69 | 0.6369 |
| 0.688681 | 35 | 94.98 | 65.00 | 94.98 | 0.5997 |
| 0.743972 | 40 | 95.95 | 60.00 | 95.95 | 0.5595 |
| 0.789471 | 45 | 96.71 | 55.00 | 96.71 | 0.5171 |
| 0.827839 | 50 | 97.31 | 50.00 | 97.31 | 0.4732 |
| 0.933922 | 75 | 99.91 | 25.00 | 99.35 | 0.2436 |
| 0.971637 | 95 | 81.85 | 5.000 | 99.91 | 0.0492 |

We further examined how well T1GRS discriminates T1D from non-diabetes at various thresholds compared to previous T1D GRS (**Fig. 3D**)^18,20^. In previous T1D GRS, the 50^th^ T1D percentile (50% sensitivity) corresponds to a false positive rate of around 3.3-6%, and at this same sensitivity T1GRS has a lower false positive rate (2.3%)^18,20^. Similarly, at a false positive rate of 5%, the sensitivity of T1GRS is 65% which is a 30% increase in the number of accurately classified T1D individuals at this threshold compared to previous GRS^20^. At the 75^th^ T1D percentile (25% sensitivity) the false positive rate is also reduced (0.65%) compared to previous GRS, and only 128 controls fall above this threshold. The lower end of prediction range is improved as well, where the 5^th^ percentile (95% sensitivity) for T1D in T1GRS had a higher specificity of 71.83% compared to previous GRS^18,20^.

We next examined differences at the MHC locus in classifying T1D in T1GRS compared to GRS2. The GRS2 model assigns specific MHC alleles based on proxy variants in an individual which in some cases can lead to incompatible MHC class II allele configurations (i.e. more than two alleles). In our samples, a total of 978 individuals (630 T1D, 348 non-diabetic) had incompatible MHC class II allele status using GRS2. By comparison, T1GRS directly uses the variants imputed from the HLA reference panel and thus avoids these incompatibilities. Of the 630 T1D individuals with incompatible MHC class II alleles in GRS2, 70.0% (441) were classified above the 50^th^ percentile in T1GRS (**Fig. 3E**). We further imputed HLA haplotypes in our samples using the SNP2HLA T1DGC reference panel as this contained a variant HLA-DQB1*02:02 necessary to fully characterize DR3/DR4 status^32^. We identified 3,810 individuals heterozygous for high-risk DR3/DR4 alleles of which 3,290 had T1D. A relatively large proportion (8.8%, 290) of the T1D individuals heterozygous for DR3/DR4 have HLA-DQ allele ambiguity in GRS2, whereas 90.7% of these individuals are classified above the 50^th^ percentile in T1GRS. (**Fig. 3E**).

We next determined the impact of variants outside the MHC locus on prediction of T1D. Of the 4,613 T1D individuals highly predicted by T1GRS (above the 50th T1D percentile) and without ambiguous HLA alleles in GRS2, 1,282 (27.8%) fell below the published GRS2 50th T1D percentile (14.6). (**Supplementary Fig. 4A**). Of these 1,282 T1D samples, 21.1% shifted above the 50^th^ T1D percentile in the non-MHC model using T1GRS, suggesting that T1GRS had improved capture of non-MHC risk for these individuals. Furthermore, the average non-MHC percentile of all 4,613 T1D samples was significantly higher in T1GRS compared to GRS2 (59.63% vs. 49.98%, paired t-test P=1.56×10^−126^) (**Fig. 3F**). Risk prediction in T1GRS was augmented by the inclusion of 60 additional genome-wide loci compared to previous scores, several of which are rare variants with relatively large effects (odds ratio [OR]>2) on T1D and may impact predictions for individuals carrying rare alleles. For example, the *CEL* locus contains a rare T1D risk variant (OR=2.8, MAF.0015), and among T1D individuals carrying the *CEL* risk allele the non-MHC percentile was significantly higher in T1GRS compared to GRS2 (paired T-test P=5.28×10^−3^) (**Supplementary Fig. 4B**).

These results overall demonstrate that T1GRS has improved diagnostic ability compared to previous T1D GRS, which is due, in part, to improved capture of risk variant effects genome-wide.

### Genetic prediction of T1D using machine learning identifies non-linear interactions

One key benefit of a machine learning model compared to previous additive genetic risk scores is the improved ability to identify non-linear interactions between features. We determined to what extent the added predictive value of genome-wide risk in T1GRS was due to capturing interactions between variants. We computed an ‘additive’ non-MHC T1GRS model where all 102 risk alleles were weighted by T1D effect size derived from the T1D meta-analysis. In highly predicted T1D donors above the 50^th^ T1GRS percentile, the ‘additive’ T1GRS model had very similar average non-MHC risk percentile to T1D GRS2 (GRS2=49.98% vs. additive T1GRS 50.15%, paired t-test P =0.588) (**Fig. 3F**). When examining the ability of the ‘additive’ non-MHC T1GRS model to discriminate T1D from non-diabetes, the inclusion of additional T1D risk loci significantly improved over GRS2 (AUC=0.739, p=1.18×10^−26^). However, the performance of this ‘additive’ T1GRS model was in turn substantially lower than the full non-MHC T1GRS model which considers interactions (p=1.33×10^−50^) (**Supplementary Fig. 4C**). These results argue that the improved predictions in T1GRS using genome-wide variants is due to both the inclusion of additional risk variants as well as capturing interactions between variants.

Next, we identified features of T1GRS influencing the classification of T1D in individuals using Shapley (SHAP) analysis. The strongest features overall were the tag SNP for HLA-DQB1*57 (rs1064173) as well as the lead variants at the *INS* and *PTPN22* loci (**Fig. 4A**). Several alleles had extensive variability in SHAP values across individuals indicating broad interactions with other features; for example, among individuals homozygous for the risk allele of the HLA-DQB1*57 tag rs1064173 (**Fig 4A,C**). To characterize interactions between variants, we calculated Shapley interaction (SHAP int) values for each pair of variants in the all variant, MHC-only, and non-MHC variant models. We evaluated the significance of interactions using a permutation test and considered interactions significant at FDR<.05 (**see Methods**). Most permuted interactions had SHAP int scores of zero, and the top interactions in each model were all significantly larger than expected (**Supplementary Tables 11,12; Supplementary Fig.5**). The strongest interaction was between HLA-DQB1*57 (rs1064173) and HLA-DRB1*13 (rs1391373) (SHAP int=1234.73) which reflects the established interaction resulting from HLA-DR3 and -DR4 haplotypes (**Fig. 4A,B**) ^25^. HLA-DQB1*57 significantly interacted with other HLA alleles including a DR3-DQ2 allele (HLA-DQB1*02:01 SHAP int=976.09), the protective DRB1*15:01/DQB1*06:02 haplotype (rs9268652 SHAP int=1105.88), and the novel non-coding signal rs9276235 near *HLA-DRB1/HLA-DQA2* (SHAP int=841.29) (**Fig. 4B,D**). The non-coding signal rs9276235 also significantly interacted with many other signals across the HLA locus (**Fig 4D**).

**Figure 4.**
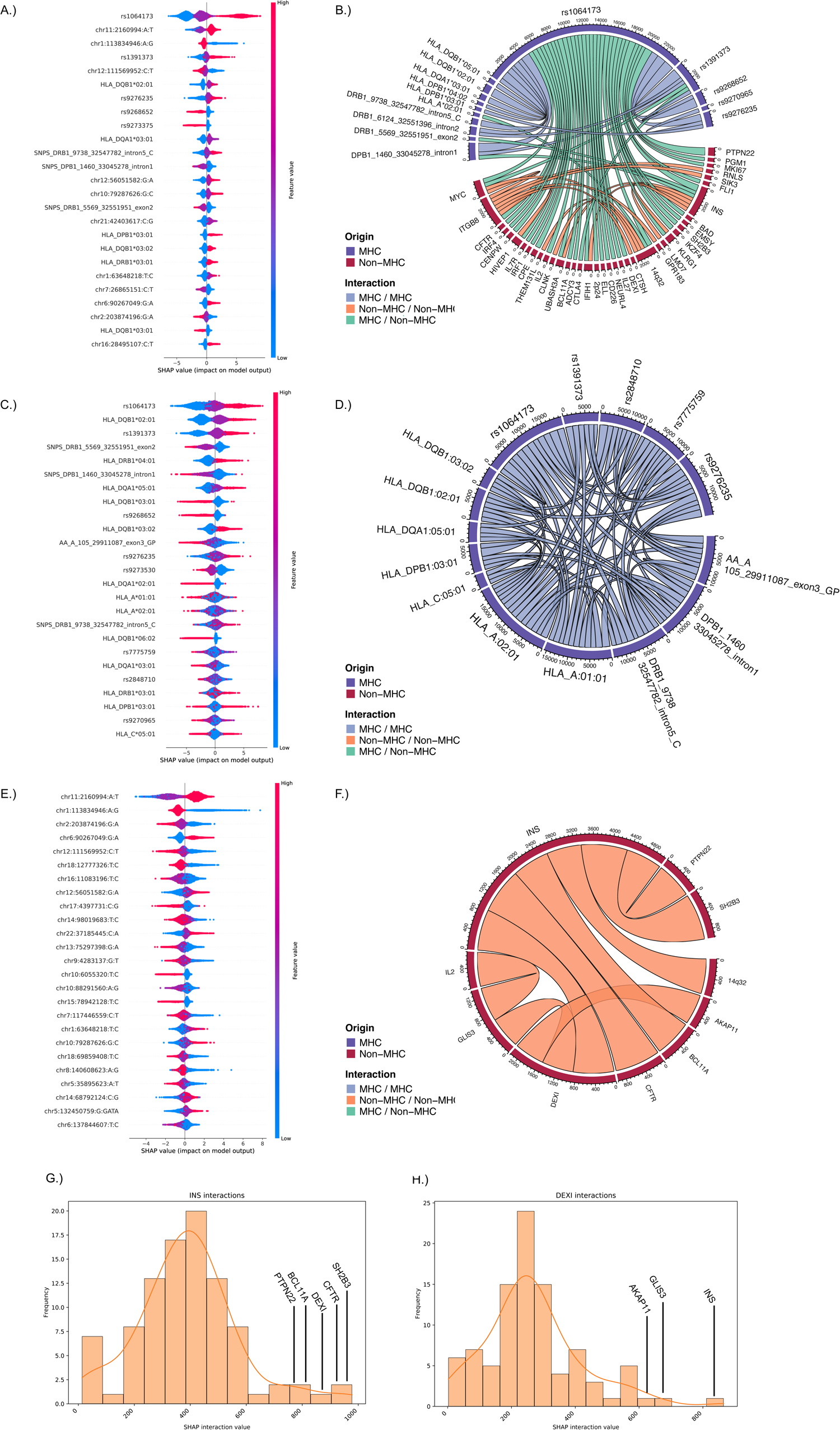
Feature importance and interactions in T1GRS. SHAP analysis results of feature importance in the top 25 features for each model (left). Red, purple and blue colors represent the contribution of 2,1, or 0 copies of the alternate allele. The spread along the x-axis indicates interactions with other variants that can alter the overall SHAP value where larger SHAP values have a greater impact on the T1GRS performance. Chord diagrams map the strongest interactions within each model (right) where blue represents an MHC-MHC interaction, green is non-MHC-MHC interaction, and orange is non-MHC-non-MHC interaction. SHAP analysis results of feature importance (left) and interactions (right) in the “reduced” T1GRS model using **A,B.** all 199 MHC and non-MHC variants. **C,D.** 97 MHC-only variants and **E,F.** 102 genome-wide non-MHC signals. Below are histograms quantifying SHAP interaction strength in the non-MHC model for **G.** the *INS* locus and **H.** the *DEXI* locus.

We also identified significant interactions (FDR<0.05) between risk variants genome-wide (**Fig. 4B,E,F**). At several loci, fine-mapping resulted in different variants representing the locus, such as at *INS* where the T1GRS variant rs689 (PIP=0.865) had higher casual probability than the variant rs3842753 (PIP=0.135) included in previous GRS^18^, which likely improves the characterization of interactions involving these loci. We identified significant interactions between MHC and non-MHC loci, most notably DQB1*57 and *PTPN22* (SHAP int=934.07) and *INS* (SHAP int=793.46) (**Fig. 4B**). There were also significant interactions between pairs of non-MHC loci, which revealed synergistic relationships between variants acting both within and across cell types. Many of the strongest non-MHC interactions were between the *INS* locus and loci implicated in immune or pancreatic cell function such as *SH2B3* (SHAP int.=978.10), *CFTR* (SHAP int.=959.74), *DEXI* (SHAP int.=864.76), *BCL11A* (SHAP int.=784.40), and *PTPN22* (SHAP int.=760.50) (**Fig. 4F,G**)^10,33–36^. In addition to the *INS* locus interaction, we also identified significant interactions between the *DEXI* locus and other putative beta cell signals including *GLIS3* (SHAP int.=667.19) and *AKAP11* (SHAP int.=635.95) (**Fig. 4F,H**)^37^.

At some individuals there were large differences in the predictions of T1D in T1GRS compared to GRS2. To highlight differences in predictions between models, we examined the risk profile of a T1D individual highly predicted in T1GRS (77^th^ percentile) but not in GRS2 (23^rd^ percentile). We determined the importance of each feature to T1GRS predictions in this individual using SHAP values, where positive and negative values indicate increased and decreased risk, respectively (**Supplementary Fig. 5G-I**). The DQB1*57 tag SNP had the largest contribution to the predictions in the overall score (+5.85) and the MHC-only score (+7.88), where the magnitude of the effect indicated the presence of interactions (**Supplementary Fig. 5G,H**). By comparison, in GRS2, MHC risk is primarily determined by HLA class II alleles and in this individual HLA-DR4-DQ8 risk is mitigated by a protective DQ4.2 haplotype. The MHC percentile was higher in T1GRS (63%) compared to thus GRS2 (32%) which is likely driven by interactions with DQB1*57. In addition, this individual had large contributions from non-MHC loci including several not in previous scores *IRF4* (+1.02) and *VSIR* (+1.00) which added substantial risk to this individual (**Supplementary Fig. 5G**). The non-MHC percentile was higher in T1GRS (70^th^ percentile) than GRS2 (24^th^ percentile) as well as the ‘additive’ T1GRS model (43^rd^ percentile), again indicating that interactions are contributing to the higher prediction in T1GRS. In sum, multiple factors led to improved predictions for this individual including additional loci and interactions between loci.

### Clustering of European individuals by T1D risk reveals disease heterogeneity

We next sought to determine to what extent T1D individuals had heterogeneity in their genetic risk profiles (**see Methods**) (**Fig. 5**). In brief, we performed principal components analysis (PCA) of the SHAP features for each T1D donor from the T1GRS model and k-means clustering of the top 10 PCs. We visualized the resulting clusters using dimension reduction of the PCs with UMAP (**Fig. 5AB, Supplementary Fig. 6**).

**Figure 5.**
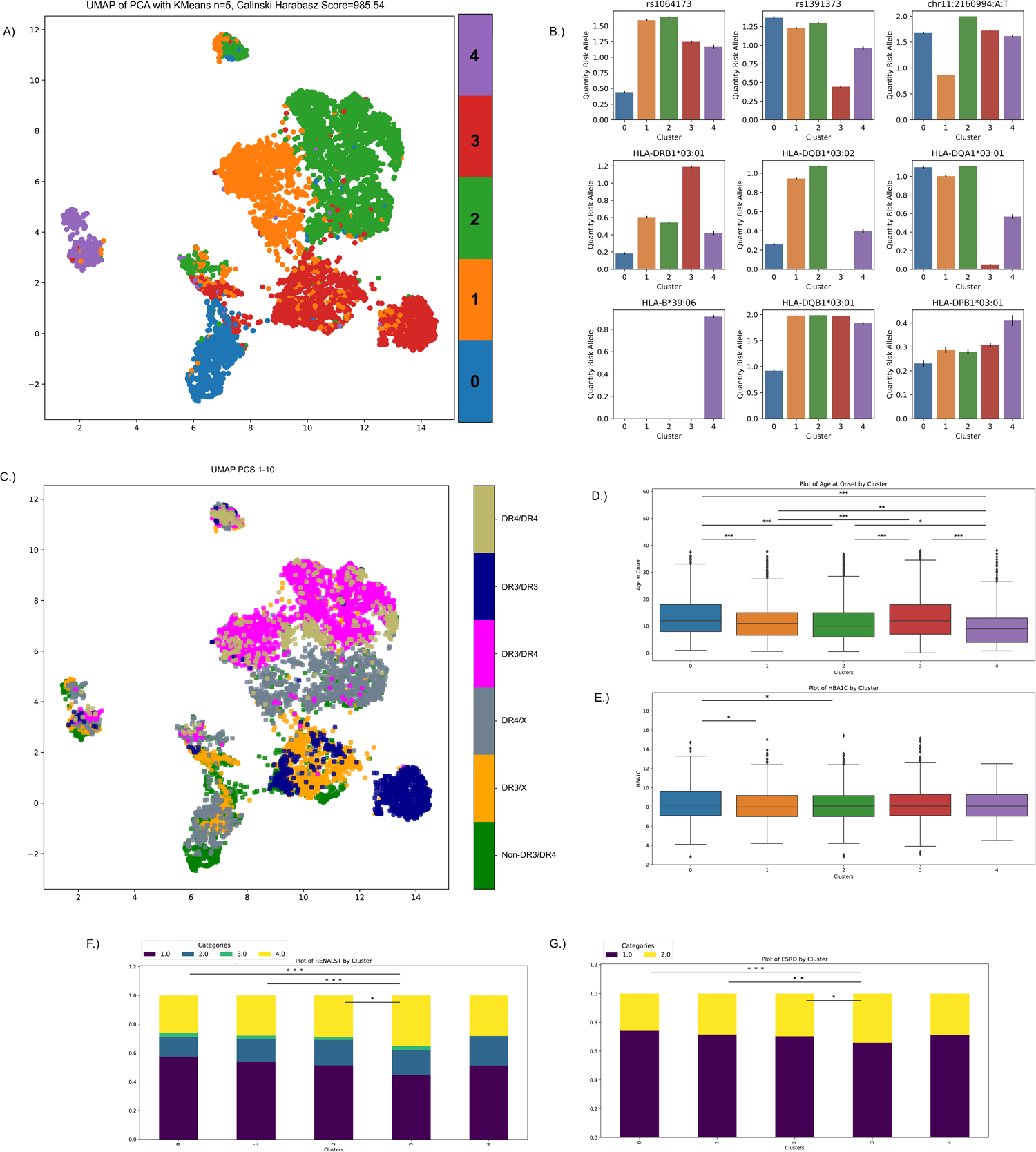
Genetic heterogeneity in T1D individuals. **A.** UMAP of the first 10 PCs showing clustering of T1D individuals defined by feature importance in T1GRS. The five clusters are separated by color. **B.** The mean quantity of risk allele contribution and standard error across the 5 clusters in 9 variants with strong feature influence in the clustering. **C.** HLA-DR3/DR4 status of T1D individuals overlayed on the UMAP plot. **D,E.** Box and whisker plots showing distributions of T1D age of disease onset and HBA1C levels at recruitment for each cluster. **F,G.** Stacked barplot showing distribution of renal function stages and end-stage renal disease for each cluster. P-values are pairwise t-tests between groups (*<0.05, **<0.01, ***<0.001).

In total we identified five clusters of T1D individuals (Calinski Harabasz Score=985.54), which were most strongly distinguished by different combinations of HLA alleles as well as the INS locus (**Fig. 5B**). Two clusters (C1, C2) are marked by the presence of the HLA-DQB1*57 amino acid and are further separated based on the presence of the INS risk allele (**Fig. 5A,B**). Cluster (C4) is described by the presence of HLA-B*39 alleles and lack of the protective HLA-DR15-DQ6 haplotype (**Fig. 5A,B,D**). Cluster (C0) is marked by the lack of HLA-DQB1*57 and the presence of HLA-DP alleles and HLA-DQB1*03:01 (**Fig. 5A,B**), and contains many of the non-MHC-DR3/DR4 samples previously shown to have distinct genetic risk (**Fig. 5C,D**) ^38^. Finally, cluster (C3) is marked by the presence of HLA-DRB1*13 and HLA-DR3 alleles. (**Fig. 5A-C**).

We finally determined whether groups of T1D individuals defined from genetic profiles had differences in clinical properties including age of onset, HbA1C level, and chronic kidney disease. We observed highly significant differences in average age of onset across groups (ANOVA P=1.1×10^−21^). For example, the C4 cluster (HLA-B*39+/ HLA-DR15-DQ6-) had significantly lower age of onset (10.6 years) than other clusters in particular compared to the C0 cluster with many non-DR3/DR4 samples (T-test P=2.30×10^−11^) and the C3 cluster (T-test P=2.90×10^−9^) (**Fig. 5C,D**). The C0 cluster also had highest HbA1C levels at recruitment (8.05%), which may indicate poorer management of symptoms in these individuals (**Fig. 5E**). We also identified a significant difference in the rate of late-stage chronic kidney disease across clusters (Kruskal-Wallis P=0.031). The C3 cluster (DRB1*13+/HLA-DR3+) exhibited a larger proportion of late-stage chronic kidney disease compared to the C0 cluster (Mann-Whitney U P=4.12×10^−3^), although they both had the latest ages of onset (**Fig. 5F**). There was similar, although not significant, difference in the rate of end-stage renal disease (ESRD) across clusters (Kruskal-Wallis P=0.10), although there was a significant difference in ESRD between C3 and C0 (Mann-Whitney U P=7.83×10^−3^).

Overall, these results reveal clusters of T1D individuals defined based on genetic risk that have evidence for heterogeneity in the clinical presentation of T1D and complications.

## Conclusions

We performed extensive genetic association and fine-mapping analysis in European ancestry individuals which revealed 165 independent risk signals for T1D, including many novel signals which provide new biological insight and potential therapeutic targets for T1D. Using these risk signals we then developed a model T1GRS to predict T1D in European ancestry individuals from genetic data which has improved accuracy compared to other contemporary risk scores. The T1GRS model in particular improved prediction of T1D outside of the MHC locus, both due to the inclusion of additional common and rare risk loci as well as by capturing interactions between loci. In addition, interactions provide potential insight into the mechanistic basis of trait association for risk loci both within and across disease-relevant cell types. Previous T1D GRS have comparatively focused more heavily on risk alleles at the HLA locus. T1GRS will therefore likely help improve prediction of T1D particularly in individuals who may not have large-effect HLA risk alleles but carry an excess burden of polygenic risk, or in individuals with complex interactions between risk alleles genome-wide not captured by additive scores. Although trained using individuals of European ancestry, T1GRS also has similar predictive ability to discriminate T1D in African Americans compared to a GRS specifically designed for this population. Future studies that train similar models using large association studies in non-European genetic individuals will undoubtedly further improve upon the prediction of T1D in African American and other ancestries.

The risk prediction model also enabled us to group T1D individuals based on genetic risk profiles, and these groups had evidence for heterogeneity in clinical features such as age of onset and severity of complications such as renal disease. In some cases, these patterns were unknown or unexpected, such as a significantly lower age of onset in HLA-B*39+/HLA-DR15-DQ6-individuals and significantly higher rate of renal complications. Recent studies have argued for the existence of distinct endotypes of T1D one of which is characterized by younger age of onset, aggressive insulitis and beta cell destruction, and abnormal proinsulin processing, and the other characterized by older age of onset, less insulitis and normal proinsulin processing^39^. These endotypes were defined using pancreatic tissue from few donors and relatively few islets per donor, however, and the extent to which these patterns generalize to the broader T1D population is unclear. Furthermore, many of these measurements are not feasible at present to collect from living donors, limiting their clinical value. Given evidence for heterogeneity in genetic risk in T1D individuals and the ability to obtain genetic profiles from all donors, genetic background should be considered in definitions of T1D heterogeneity, in combination with other biomarkers.

A primary goal of this study was to provide a predictive model of T1D that could be easily applied using genetic data from any samples of interest. Our model uses only variants present on the contemporary TOPMed r3 and Michigan HLA reference panels, where available web tools enable any user to impute genotyping microarray or sequencing data into these panels. Previous approaches have utilized proxy variants to capture HLA alleles which may not be present in current reference panels or on all genotyping microarrays. Furthermore, using proxy SNPs to phase HLA alleles leads to ambiguity in HLA typing for a proportion of individuals, including for individuals with high-risk DR3/DR3 haplotypes, complicating risk prediction in these individuals. By comparison, the T1GRS model uses variants directly from imputation panels for risk prediction and therefore avoids many issues with ambiguous allele phasing. Second, we facilitated the prediction of T1D using T1GRS by providing scripts that take variant files directly from TOPMed and Michigan HLA outputs as input to the model. These scripts and models are provided in containers which can be easily run in any computing environment.

In total, T1GRS provides a new predictive of model of T1D which should facilitate and improve the use of genetic risk scores for T1D in both basic research and clinical applications.

## Research Design and Methods

### Research subjects and genotype imputation

For the MHC analysis, we compiled genotype data from 29,746 T1D case and control individuals of European ancestry from five publicly available cohorts (**Supplementary Table 1**). T1D cases were matched to control subjects by ancestry and, where possible, genotype array as previously described^10^. We performed quality control on variants using the HRC imputation preparation program (version 4.2.9, https://www.well.ox.ac.uk/~wrayner/tools/) and PLINK to remove variants with MAF<1%, missing genotypes >5%, in violation of Hardy-Weinberg equilibrium (HWE *P* <1×10^−5^ in control cohorts and HWE *P*<1×10^−10^ in case cohorts), allele ambiguity and difference in allele frequency > 0.2 compared to HRC r1.1 reference panel^40,41^. We imputed 55,615 variants from the ‘Four-digit Multi-ethnic HLA reference panel (v1)’ from Michigan imputation server and retained variants with r^2^>0.5 and a standard deviation in control allele frequency< 0.055 to test for association in the 29,746 samples^42^.

To examine genetic risk outside of the MHC, we compiled association data from 20,355 case and 797,363 control individuals from 817,718 European ancestry cohorts matched by ancestry and genotype array where possible, as described previously^10^ (**Supplementary Table 1**). For the FinnGen cohort, we downloaded the summary statistics from the r10 version for “T1D_Early” which includes 2,832 individuals diagnosed with T1D under the age of 20 and excludes individuals diagnosed with T2D (r10.risteys.finngen.fi/endpoints/T1D_EARLY).

We used genotyping microarray data for 263 individuals in the Network for Pancreatic Organ Donors with Diabetes (nPOD) including 115 T1D cases and 148 individuals without T1D. Additionally, we used 1,999 T2D individuals from the WTCCC1 cohort^29,30^. To examine predictive ability of T1GRS in individuals of African American ancestry, we used 284 T1D samples from SEARCH consortium and 404 non-disease individuals from CLEAR cohort^43,44^. For all cohorts, we performed variant quality control as stated above prior to imputation with Michigan HLA and TOPMed imputation panels.

### Association testing and meta-analysis at the MHC locus

In the MHC locus, we tested variants for association across a 4 MB locus on chromosome 6 from 30 Mb to 34 Mb (hg19). We used firth bias corrected logistic regression in EPACTS and tested variants for association with MAF>1% including the first four genotype PCs and sex as covariates^45^. For both the MHC and genome-wide association analyses, we combined summary statistics from all five cohorts using a fixed effects inverse variance weighted meta-analysis.

### Conditional analysis of independent signals

We performed conditional forward stepwise analysis in the MHC locus by including the most significant variant from each meta-analysis as a covariate in the association tests for each cohort and then re-doing the meta-analysis. We repeated this process by iteratively adding each new variant to the model until no significant variants (p<5×10^−8^) remained in the meta-analysis.

We also performed a ‘preconditional’ analysis to examine the effect of signals outside of 70 established class I and II HLA risk alleles and 20 DR3/DR4 pairwise HLA haplotypic and allelic interactions^25,46–48^. We added 90 covariates to our model described above to capture the additive effect of each alternate allele for 70 HLA risk alleles and a binary column for each of 20 interactions prior to performing the association tests and meta-analysis. We then performed conditional forward stepwise regression iteratively adding the most significant variant to the model and re-performing the meta-analysis until no variants reached p<5×10^−8^.

### Credible set generation for signals in the MHC

For all conditional signals identified in the MHC locus though stepwise analysis, we generated 95% credible sets (CSs). We first identified any variants with linkage to the lead variant in each signal (r^2^>0.1) and we calculated Bayes Factor for each variant in this set based on the effect and standard error as described in the method of Wakefield^49^. We then generated posterior inclusion probability (PIP) scores by dividing the Bayes Factor by the total sum of the Bayes Factors for all variants in the set. We ordered variants in descending order and included variants up to the 95% threshold in each credible set.

### SuSIE fine-mapping of the non-MHC loci

We used SuSIE to fine-map significant non-MHC loci identified through two rounds of variant clumping using PLINK (’--clump-p1 5e-8 --clump-p2 0.05 --clump-r2 0.1 --clump-kb 10000’; ‘--clump-p1 5e-8 --clump-r2 0 --clump-kb 500’). We then generated loci 500kb around the variants included in each clumped region. We identified 92 genome-wide significant loci and included 10 additional loci not reaching genome wide significance that were previously reported (*CDKN1C*, *CYP27B1*, *LMO7*, *CCR7*, *17q24*, *ACOXL*, *CCR5*, *IRF2*, *TAGAP*, and *6q27*) for a total of 102 loci. We created 95% credible sets for each locus in SuSIE using genotypes from 6 dbGAP cohorts including 32,518 samples (DCCT, GENIE-ROI, GENIE-UK, GoKIND, T1DGC, and WTCCC1) to define the LD matrix and setting parameters to ‘L=10, coverage=0.95, min_abs_corr=0.01, max_iter=50000’. For complex loci with multiple signals identified in a previous study (*DLK1*, *IFIH1*, *TYK2*, *IL10*, *PTPN2*, *AIRE*, *UBASH3A*, *CTLA4*, and *IL2RA*), we re-computed the T1D meta-analysis using only the 6 dbGAP cohorts with genotype data included in the LD matrix listed above ^10^. Lead variants were defined as the variant with the largest posterior inclusion probability (PIP) for the signal. We defined novel loci as variants that reached genome wide significance and mapped 500kb away from any other established loci.

### Annotations of credible sets

We leveraged genomic datasets^27,50^ and examined preferential transcription factor (TF) binding motifs to conduct a functional analysis in novel credible sets. We overlapped all variants in novel credible sets with accessible chromatin peaks in 46 immune and 12 pancreatic cell types^27,50^. Then we tested each variant for preferential allelic binding to transcription factor motifs using FIMO^51^. We also leveraged databases such as GTEx and JASPAR to annotate credible set variants.

### Constructing a non-linear Machine learning polygenic risk score

We leveraged 199 variants including the lead variant from 27 independent MHC signals, 70 additional established HLA associated alleles, and 102 non-MHC most probable variants identified using SuSIE. We developed two models based on the XGBoost classifier framework^52,53^, one with the 199 variants alone (“variant only” model), and another with additional covariates of sex, PC1-4, and binary covariates for each cohort (“full” model). This approach generates a probability from zero to one that a sample has T1D which can be treated as a genetic risk score. Five cohorts containing 10,107 T1D and 19,639 non-diabetes samples were combined into one genotype matrix which was then randomly split into 10 subsections for cross fold validation. Over 10 iterations, a model was trained on 90% of the data and evaluated on the remaining 10%. The probability scores for each sample in each testing fold were recorded and used for the overall AUC. This process was identical in the T1GRS evaluation conditions: full (199 variants), MHC only (97 variants), and non-MHC only (102 variants) models. A representative model for each of the evaluation conditions was trained on all samples and applied to the nPOD validation cohort of 115 T1D and 148 non-diabetes samples. A standard random seed was set to ensure reproducibility and identical hyperparameters were used for each version of the model. Additionally, versions of each model were trained without PC and cohort information (“variant only” models) and evaluated in an identical manner.

### Feature importance and interaction analysis

Feature importance and interaction within non-linear models were calculated using the SHAP machine learning interpretability suite (https://shap.readthedocs.io/en/latest/)^54^. SHAP, which stands for SHapley Additive exPlanations, is a unified approach to explain the output of any machine learning model. It is based on cooperative game theory and the concept of Shapley values. SHAP values assign each feature an importance value for a particular prediction in the context of a specific model. These values allow for non-linear interactions between features to be accounted for on a per-individual basis and enables ranking pairwise feature interactions by magnitude. Each model was run through the standard SHAP python pipeline and the feature importance was recorded. Feature interaction analysis was performed using the shap_interaction_values function.

### PCA analysis and kmeans clustering of SHAP values

SHAP analysis results in a vector of patient specific feature contributions to outcome, where the same feature can have different contributions to outcome in different patients, based on the additive non-linear effects described by SHAP analysis. To uncover differences in the etiologies of phenotypically similar disease we performed PCA analysis on these SHAP feature contribution vectors in T1D samples, then created a UMAP using the top 20 PCs from PCA analysis and n_neighbors=10. Next, we performed unsupervised kmeans clustering on the top 10 PCs. A range of clusters were tested from k=2-10 and we found that k=5 resulted in the highest Calinski Harabasz Score (**Fig. 5A**), indicating that k=5 clusters yielded the greatest separation between clusters. Clinical features including age of onset, HbA1C (%), chronic kidney disease (RENALST stages 1-4), and end stage renal disease ESRD (1 vs 2) were compared between clusters using pairwise t-tests. An ANOVA was used to compare age of onset between clusters while Kruskal-Wallis tests were applied to the other phenotypes.

### Statistical analysis of the genetic risk score

We then calculated GRS2 using 60 exact TOPMed variants, two exact Michigan HLA for rs116522341, rs1281934, and the proxy variants DQB1*06:02, B*18:01, DPB1*03:01, rs1611547, and rs114170382 from Michigan HLA for rs17843689, rs371250843, rs559242105, rs144530872, and rs149663102 respectively. In GRS2, we excluded individuals with more than 2 HLA-DR/DQ proxy SNPs according to the published methods^18^. Within each GRS, we examined the total genetic risk score, and its components of MHC (Michigan HLA) and non-MHC variants (TOPMed). We calculated the area under the curve (AUC) for the receiver operator characteristic (ROC) analysis to assess the differentiation power of each GRS for T1D. We then tested the difference between each AUC using the deLong test. First, we compared the T1GRS model to GRS2 in individuals with T1D and those without in the “full” model and in the “variant only” model. Next, we validated T1GRS using an independent set of individuals in the nPOD biorepository to differentiate between T1D and individuals without disease and using the T1D from nPOD and 1,999 Type 2 diabetes (T2D) from WTCCC1.

We also calculated a published African American risk score in 284 T1D and 404 non-diabetes samples from SEARCH and CLEAR studies to compare to T1GRS^31,43,44^. We used TOPMed to impute rs34850435, rs9271594, rs9273363, rs2290400, rs689, Michigan HLA to impute rs2187668, and used rs9268838 as a proxy for rs34303755 (r^2^=0.849, D’=1.0 in African Americans).

Lastly, we generated a scale for T1GRS scores using the number of individuals with T1D who fell at various percentiles and calculated a diagnostic for each GRS value using the Youden index (sensitivity + specificity −1). We calculated sensitivity at each GRS score on the scale as TP/(TP + FN) and specificity as TN/(TN+FP)^55^. Additionally, we defined DR3/DR4 individuals using four-digit HLA alleles imputed from the T1DGC reference panel using SNP2HLA^32^. DR3 status was classified by the presence of HLA-DRB1*03:01-DQB1*02:01, while DR4 was identified by the presence of HLA-DRB1*04:01/02/04/05/08-DQB1*03:02/04/02:02^56^.

## Code availability

The T1GRS model and code to generate T1D predictions using T1GRS is available at https://github.com/Gaulton-Lab/t1grs

## Data availability

Summary statistics from the T1D GWAS will be made available in the GWAS catalog.

## Author contributions

K.J.G. and H.C. supervised the work in this study. K.J.G., H.C., C.M. and T.S. wrote the manuscript. C.M. performed all genetic association analyses, contributed to the development of the predictive model, and evaluated model performance and other downstream analyses. T.S. developed and trained the predictive model and performed evaluations of model performance and other downstream analyses. P.K. developed the code repository and containerization scripts.

## Acknowledgements

The work in this study was funded by UCSD internal funding including through the Winkler Endowed Chair in Type 1 Diabetes to K.J.G., and the Mark Foundation for Cancer Research to H.C. KJG has done consulting for Genentech, received honoraria from Pfizer, and is a shareholder of Neurocrine biosciences.

## DCCT/EDIC

The Diabetes Control and Complications Trial (DCCT) and its follow-up the Epidemiology of Diabetes Interventions and Complications (EDIC) study were conducted by the DCCT/EDIC Research Group and supported by National Institute of Health grants and contracts and by the General Clinical Research Center Program, NCRR. The data from the DCCT/EDIC study were supplied by the NIDDK Central Repositories.

## GENIE

The Genetics of Nephropathy, an International Effort (GENIE) study was conducted by the GENIE Investigators and supported by the National Institute of Diabetes and Digestive and Kidney Diseases (NIDDK). The data from the GENIE study reported here were supplied by the GENIE investigators from the Broad Institute of MIT and Harvard, Queens University Belfast and the University of Dublin.

## GoKinD

The Genetics of Kidneys in Diabetes (GoKinD) Study was conducted by the GoKinD Investigators and supported by the Juvenile Diabetes Research Foundation, the CDC, and the Special Statutory Funding Program for Type 1 Diabetes Research administered by the National Institute of Diabetes and Digestive and Kidney Diseases (NIDDK). The data [and samples] from the GoKinD study were supplied by the NIDDK Central Repositories. This manuscript was not prepared in collaboration with Investigators of the GoKinD study and does not necessarily reflect the opinions or views of the GoKinD study, the NIDDK Central Repositories, or the NIDDK.

## T1DGC

This research utilizes resources provided by the Type 1 Diabetes Genetics Consortium (T1DGC), a collaborative clinical study sponsored by the National Institute of Diabetes and Digestive and Kidney Diseases (NIDDK), National Institute of Allergy and Infectious Diseases (NIAID), National Human Genome Research Institute (NHGRI), National Institute of Child Health and Human Development (NICHD), and the Juvenile Diabetes Research Foundation International (JDRF) and supported by U01 DK062418. The UK case series collection was additionally funded by the JDRF and Wellcome Trust and the National Institute for Health Research Cambridge Biomedical Centre, at the Cambridge Institute for Medical Research, UK (CIMR), which is in receipt of a Wellcome Trust Strategic Award (079895). The data from the T1DGC study were supplied by dbGAP. This manuscript was not prepared in collaboration with Investigators of the T1DGC study and does not necessarily reflect the opinions or views of the T1DGC study or the study sponsors.

## T1DGC (ASP/UK GRID)

This research was performed under the auspices of the Type 1 Diabetes Genetics Consortium, a collaborative clinical study sponsored by the National Institute of Diabetes and Digestive and Kidney Diseases (NIDDK), National Institute of Allergy and Infectious Diseases (NIAID), National Human Genome Research Institute (NHGRI), National Institute of Child Health and Human Development (NICHD), and Juvenile Diabetes Research Foundation International (JDRF).

## WTCCC

This study makes use of data generated by the Wellcome Trust Case Control Consortium. A full list of the investigators who contributed to the generation of the data is available from www.wtccc.org.uk. Funding for the project was provided by the Wellcome Trust under award 076113.

## UK Biobank

Data from the UK Biobank was accessed under application 24058.

## FinnGen

The FinnGen study is a large-scale genomics initiative that has analyzed over 500,000 Finnish biobank samples and correlated genetic variation with health data to understand disease mechanisms and predispositions. The project is a collaboration between research organisations and biobanks within Finland and international industry partners. We want to acknowledge the participants and investigators of the FinnGen study.

## CLEAR

The data used for the analyses described in this paper were obtained from the database of Genotypes and Phenotypes (dbGaP), at http://www.ncbi.nlm.nih.gov/gap. Genotype and phenotype data for the study “Genome-Wide Association Study in African-Americans with Rheumatoid Arthritis” were provided by Dr. S. Louis Bridges, Jr. University of Alabama at Birmingham. The GWAS study (R01AR057202) was supported by awards from the National Institute of Arthritis and Musculoskeletal and Skin Diseases. The cohorts used in the GWAS study were derived from participants in the Consortium for the Longitudinal Evaluation of African-Americans with Early Rheumatoid Arthritis (CLEAR I and II) case-control observational studies, supported by a research contract awarded by the National Institute of Arthritis and Musculoskeletal and Skin Diseases. The consortium is a collaborative effort among five academic institutions: University of Alabama at Birmingham, Birmingham, AL (Coordinating Center); Grady Hospital/Emory University, Atlanta, GA; University of North Carolina, Chapel Hill, NC; Medical University of South Carolina, SC; and Washington University, St. Louis, MO. For specific publication describing the CLEAR studies, CLEAR collaborators, see https://www.uab.edu/medicine/rheumatology/research/70-clear and https://www.ncbi.nlm.nih.gov/pmc/articles/PMC3052790/.

## SEARCH

The SEARCH for Diabetes in Youth study was conducted by the SEARCH Investigators and supported by the National Institute of Diabetes and Digestive and Kidney Diseases (NIDDK). The data from the SEARCH Study reported here were supplied by the institutions (funding) listed below: Kaiser Permanente Southern California (U18DP006133, U48/CCU919219, U01 DP000246, and U18DP002714), University of Colorado Denver (U18DP006139, U48/CCU819241-3, U01 DP000247, and U18DP000247-06A1), Cincinnati’s Children’s Hospital Medical Center (U18DP006134, U48/CCU519239, U01 DP000248, and 1U18DP002709), University of North Carolina at Chapel Hill (U18DP006138, U48/CCU419249, U01 DP000254, and U18DP002708), Seattle Children’s Hospital (U18DP006136, U58/CCU019235-4, U01 DP000244, and U18DP002710-01), Wake Forest School of Medicine (U18DP006131, U48/CCU919219, U01 DP000250, 200-2010-35171, DP15-0020301SUPP17, and 1UC4DK108173).

## Network for Pancreatic Organ Donors with Diabetes (nPOD)

This research was performed with the support of the Network for Pancreatic Organ donors with Diabetes (nPOD; RRID:SCR_014641), a collaborative type 1 diabetes research project supported by JDRF (nPOD: 5-SRA-2018-557-Q-R) and The Leona M. & Harry B. Helmsley Charitable Trust (Grant#2018PG-T1D053). The content and views expressed are the responsibility of the authors and do not necessarily reflect the official view of nPOD. Organ Procurement Organizations (OPO) partnering with nPOD to provide research resources are listed at https://npod.org/for-partners/npod-partners/.

## CSGNM

We thank the participants of the Trinity Student Study. This study was supported by the Intramural Research Programs of the National Institutes of Health, the National Human Genome Research Institute, and the Eunice Kennedy Shriver National Institute of Child Health and Development.

## NIMH Schizophrenia Controls

Funding support for the Genome-Wide Association of Schizophrenia Study was provided by the National Institute of Mental Health (R01 MH67257, R01 MH59588, R01 MH59571, R01 MH59565, R01 MH59587, R01 MH60870, R01 MH59566, R01 MH59586, R01 MH61675, R01 MH60879, R01 MH81800, U01 MH46276, U01 MH46289 U01 MH46318, U01 MH79469, and U01 MH79470) and the genotyping of samples was provided through the Genetic Association Information Network (GAIN). The datasets used for the analyses described in this manuscript were obtained from the database of Genotypes and Phenotypes (dbGaP) found at http://www.ncbi.nlm.nih.gov/gap through dbGaP accession number phs000021.v3.p2. Samples and associated phenotype data for the Genome-Wide Association of Schizophrenia Study were provided by the Molecular Genetics of Schizophrenia Collaboration (PI: Pablo V. Gejman, Evanston Northwestern Healthcare (ENH) and Northwestern University, Evanston, IL, USA).

## Neurodevelopmental Genomics

Support for the collection of the data for Philadelphia Neurodevelopment Cohort (PNC) was provided by grant RC2MH089983 awarded to Raquel Gur and RC2MH089924 awarded to Hakon Hakonarson. Subjects were recruited and genotyped through the Center for Applied Genomics (CAG) at The Children’s Hospital in Philadelphia (CHOP). Phenotypic data collection occurred at the CAG/CHOP and at the Brain Behavior Laboratory, University of Pennsylvania.

## eMERGE Network

Group Health Cooperative/University of Washington – Funding support for Alzheimer’s Disease Patient Registry (ADPR) and Adult Changes in Thought (ACT) study was provided by a U01 from the National Institute on Aging (Eric B. Larson, PI, U01AG006781). A gift from the 3M Corporation was used to expand the ACT cohort. DNA aliquots sufficient for GWAS from ADPR Probable AD cases, who had been enrolled in Genetic Differences in Alzheimer’s Cases and Controls (Walter Kukull, PI, R01 AG007584) and obtained under that grant, were made available to eMERGE without charge. Funding support for genotyping, which was performed at Johns Hopkins University, was provided by the NIH (U01HG004438). Genome-wide association analyses were supported through a Cooperative Agreement from the National Human Genome Research Institute, U01HG004610 (Eric B. Larson, PI). Mayo Clinic – Samples and associated genotype and phenotype data used in this study were provided by the Mayo Clinic. Funding support for the Mayo Clinic was provided through a cooperative agreement with the National Human Genome Research Institute (NHGRI), Grant #: UOIHG004599; and by grant HL75794 from the National Heart Lung and Blood Institute (NHLBI). Funding support for genotyping, which was performed at The Broad Institute, was provided by the NIH (U01HG004424). Marshfield Clinic Research Foundation – Funding support for the Personalized Medicine Research Project (PMRP) was provided through a cooperative agreement (U01HG004608) with the National Human Genome Research Institute (NHGRI), with additional funding from the National Institute for General Medical Sciences (NIGMS) The samples used for PMRP analyses were obtained with funding from Marshfield Clinic, Health Resources Service Administration Office of Rural Health Policy grant number D1A RH00025, and Wisconsin Department of Commerce Technology Development Fund contract number TDF FYO10718. Funding support for genotyping, which was performed at Johns Hopkins University, was provided by the NIH (U01HG004438). Northwestern University – Samples and data used in this study were provided by the NUgene Project (www.nugene.org). Funding support for the NUgene Project was provided by the Northwestern University’s Center for Genetic Medicine, Northwestern University, and Northwestern Memorial Hospital. Assistance with phenotype harmonization was provided by the eMERGE Coordinating Center (Grant number U01HG04603). This study was funded through the NIH, NHGRI eMERGE Network (U01HG004609). Funding support for genotyping, which was performed at The Broad Institute, was provided by the NIH (U01HG004424).

## Vanderbilt University

Funding support for the Vanderbilt Genome-Electronic Records (VGER) project was provided through a cooperative agreement (U01HG004603) with the National Human Genome Research Institute (NHGRI) with additional funding from the National Institute of General Medical Sciences (NIGMS). The dataset and samples used for the VGER analyses were obtained from Vanderbilt University Medical Center’s BioVU, which is supported by institutional funding and by the Vanderbilt CTSA grant UL1RR024975 from NCRR/NIH. Funding support for genotyping, which was performed at The Broad Institute, was provided by the NIH (U01HG004424). Assistance with phenotype harmonization and genotype data cleaning was provided by the eMERGE Administrative Coordinating Center (U01HG004603) and the National Center for Biotechnology Information (NCBI). The datasets used for the analyses described in this manuscript were obtained from dbGaP at http://www.ncbi.nlm.nih.gov/gap through dbGaP accession number phs000360.v3.p1.

This manuscript was not prepared in collaboration with investigators of these studies and does not necessarily reflect the opinions or views of the DCCT/EDIC, GENIE, GoKinD, T1DGC, WTCCC, CLEAR, SEARCH studies or study groups, the NIDDK Central Repositories, the NIH, or the study sponsors.

---

**Supplementary Figure 1. Annotation of novel T1D signals at the MHC locus.** A. Locus plot showing a novel non-coding signal at the MHC locus with index variant rs9276235. The bottom of the plot shows expression QTLs for *TAP2* at this variant in GTEx for multiple tissues. B. Locus and credible set plots for novel signals identified in pre-conditioned HLA T1D association analysis. C. Annotation of a novel T1D signal upstream of *HLA-A* where variant rs7763052 maps in accessible chromatin sites for stimulated (S) and unstimulated (U) conditions of different immune cell types.

**Supplementary Figure 2: Precision-Recall Curves in T1GRS and GRS2.** Precision-Recall curves highlighting the Average Precision (AP) in purple for T1GRS and green for GRS2 in **A.** total scores **B.** MHC only scores and **C.** non-MHC scores.

**Supplementary Figure 3.** ROC statistics for T1GRS and T1D GRS2. The AUC for T1GRS is purple while the existing GRS2 is green. P-values comparing the predictive ability of GRSs are calculated using the de-Long test. The AUCs for the “variants-only” T1GRS and T1D GRS2 are compared in T1D and non-disease individuals in the discovery group of 29,746 samples for **A.** All variants **B.** MHC-only variants and **C.** non-MHC genome-wide variants. The AUCs for T1GRS and T1D GRS2 are compared in T1D and non-disease individuals from the independent nPOD test group for **D.** All variants **E.** MHC-only variants and **F.** non-MHC genome-wide variants. The AUCs for T1GRS and GRS2 are compared in Type 1 diabetes cases from the nPOD test group and Type 2 diabetes individuals from WTCCC1 for **G.** Total scores **H.** MHC variants and **I.** non-MHC variants**. J.** AUCs in T1GRS between African Americans and individuals of European ancestry with and without T1D. **K.** AUCs in African American individuals with and without diabetes in T1GRS and a 7 variant African American GRS.

**Supplementary Figure 4. A.** T1D GRS2 scores for individuals who fall above the 50^th^ T1D percentile in T1GRS. **B.** GRS2 and T1GRS non-MHC percentiles for 157 T1D individuals with a large effect rare variant (CEL). **C.** ROC statistics for non-MHC differentiation in T1D and non-disease for T1GRS ML model, T1GRS additive model, and GRS2.

**Supplementary Figure 5.** Distribution and top variant interaction SHAP values in T1GRS. **A.** 199 variant total model distribution of SHAP interactions with **B.** the top 25 interactions in the 199 variant model. **C.** 97 variant MHC only model distribution of SHAP interactions with **D.** the top 25 interactions in the 97 MHC variant model. **E.** 102 variant non-MHC only model distribution of SHAP interactions with **F.** the top 25 interactions in the 97 MHC variant model. SHAP values in a high-risk individual with T1D in **G.** 199 variant total model **H**. 97 variant MHC only model and **I**. 102 variant non-MHC only model.

**Supplementary Figure 6.** PC influences in the Cluster analysis of T1D samples. **A-J.** On the left, UMAPs of PCs 1 through 10 after kmeans (n=5) clustering T1D samples. The color variable represents the presence of a PC in a region. On the right, bar plots highlight the features with the strongest influences in each PC with larger values.

## Notes

### Author Declarations

The Institutional Review Board (IRB) of the University of California San Diego gave ethical approval for this work

## References

1 Katsarou A, Gudbjörnsdottir S, Rawshani A, et al. Type 1 diabetes mellitus. Nat Rev Dis Primer 2017; 3: 1–17.

2 Thomas NJ, Jones SE, Weedon MN, Shields BM, Oram RA, Hattersley AT. Frequency and phenotype of type 1 diabetes in the first six decades of life: a cross-sectional, genetically stratified survival analysis from UK Biobank. Lancet Diabetes Endocrinol 2018; 6: 122–9.

3 Cameron FJ, Scratch SE, Nadebaum C, et al. Neurological consequences of diabetic ketoacidosis at initial presentation of type 1 diabetes in a prospective cohort study of children. Diabetes Care 2014; 37: 1554–62.

4 Duca LM, Wang B, Rewers M, Rewers A. Diabetic Ketoacidosis at Diagnosis of Type 1 Diabetes Predicts Poor Long-term Glycemic Control. Diabetes Care 2017; 40: 1249–55.

5 Herold KC, Bundy BN, Long SA, et al. An Anti-CD3 Antibody, Teplizumab, in Relatives at Risk for Type 1 Diabetes. N Engl J Med 2019; 381: 603–13.

6 Ziegler A-G, Nepom GT. Prediction and Pathogenesis in Type 1 Diabetes. Immunity 2010; 32: 468–78.

7 Sørgjerd EP, Thorsby PM, Torjesen PA, Skorpen F, Kvaløy K, Grill V. Presence of anti-GAD in a non-diabetic population of adults; time dynamics and clinical influence: results from the HUNT study. BMJ Open Diabetes Res Care 2015; 3: e000076.

8 Noble JA, Valdes AM. Genetics of the HLA region in the prediction of type 1 diabetes. Curr Diab Rep 2011; 11: 533–42.

9 Noble JA, Valdes AM, Cook M, Klitz W, Thomson G, Erlich HA. The role of HLA class II genes in insulin-dependent diabetes mellitus: molecular analysis of 180 Caucasian, multiplex families. Am J Hum Genet 1996; 59: 1134–48.

10 Chiou J, Geusz RJ, Okino ML, et al. Interpreting type 1 diabetes risk with genetics and single-cell epigenomics. Nat 2021 5947863 2021; 594: 398–402.

11 Barrett JC, Clayton DG, Concannon P, et al. Genome-wide association study and meta-analysis find that over 40 loci affect risk of type 1 diabetes. Nat Genet 2009; 41: 703–7.

12 Bradfield JP, Qu H-Q, Wang K, et al. A genome-wide meta-analysis of six type 1 diabetes cohorts identifies multiple associated loci. PLoS Genet 2011; 7: e1002293.

13 Onengut-Gumuscu S, Chen WM, Burren O, et al. Fine mapping of type 1 diabetes susceptibility loci and evidence for colocalization of causal variants with lymphoid gene enhancers. Nat Genet 2015; 47: 381–6.

14 Grant SFA, Wells AD, Rich SS. Next steps in the identification of gene targets for type 1 diabetes. Diabetologia 2020; 63: 2260–9.

15 Uffelmann E, Huang QQ, Munung NS, et al. Genome-wide association studies. Nat Rev Methods Primer 2021; 1: 1–21.

16 Winkler C, Krumsiek J, Buettner F, et al. Feature ranking of type 1 diabetes susceptibility genes improves prediction of type 1 diabetes. Diabetologia 2014; 57: 2521–9.

17 Mishra R, Åkerlund M, Cousminer DL, et al. Genetic Discrimination Between LADA and Childhood-Onset Type 1 Diabetes Within the MHC. Diabetes Care 2019; 43: 418–25.

18 Sharp SA, Rich SS, Wood AR, et al. Development and standardization of an improved type 1 diabetes genetic risk score for use in newborn screening and incident diagnosis. In: Diabetes Care. American Diabetes Association Inc., 2019: 200–7.

19 Joglekar MV, Kaur S, Pociot F, Hardikar AA. Prediction of progression to type 1 diabetes with dynamic biomarkers and risk scores. Lancet Diabetes Endocrinol 2024; 12: 483–92.

20 Oram RA, Patel K, Hill A, et al. A Type 1 Diabetes Genetic Risk Score Can Aid Discrimination Between Type 1 and Type 2 Diabetes in Young Adults. Diabetes Care 2016; 39: 337–44.

21 Alghamdi TA, Krentz NAJ, Smith N, et al. Zmiz1 is required for mature β-cell function and mass expansion upon high fat feeding. Mol Metab 2022; 66: 101621.

22 Taheri M, Eghtedarian R, Dinger ME, Ghafouri-Fard S. Emerging roles of non-coding RNAs in the pathogenesis of type 1 diabetes mellitus. Biomed Pharmacother 2020; 129: 110509.

23 Luo Y, Kanai M, Choi W, et al. A high-resolution HLA reference panel capturing global population diversity enables multi-ethnic fine-mapping in HIV host response. Jacques Fellay; 14. DOI:10.1101/2020.07.16.20155606.

24 Todd JA, Bell JI, McDevitt HO. HLA-DQ beta gene contributes to susceptibility and resistance to insulin-dependent diabetes mellitus. Nature 1987; 329: 599–604.

25 Hu X, Deutsch AJ, Lenz TL, et al. Additive and interaction effects at three amino acid positions in HLA-DQ and HLA-DR molecules drive type 1 diabetes risk. Nat Genet 2015; 47: 898–905.

26 Zhao LP, Papadopoulos GK, Lybrand TP, et al. The KAG motif of HLA-DRB1 (β71, β74, β86) predicts seroconversion and development of type 1 diabetes. EBioMedicine 2021; 69: 103431.

27 Calderon D, Nguyen MLT, Mezger A, et al. Landscape of stimulation-responsive chromatin across diverse human immune cells. Nat Genet 2019; 51: 1494–505.

28 Yan J, Qiu Y, Ribeiro dos Santos AM, et al. Systematic analysis of binding of transcription factors to noncoding variants. Nature 2021; 591: 147–51.

29 Campbell-Thompson M, Wasserfall C, Kaddis J, et al. Network for Pancreatic Organ Donors with Diabetes (nPOD): Developing a Tissue Biobank for Type 1 Diabetes. Diabetes Metab Res Rev 2012; 28: 608–17.

30 Wellcome Trust Case Control Consortium. Genome-wide association study of 14,000 cases of seven common diseases and 3,000 shared controls. Nature 2007; 447: 661–78.

31 Onengut-Gumuscu S, Chen WM, Robertson CC, et al. Type 1 diabetes risk in African-ancestry participants and utility of an ancestry-specific genetic risk score. Diabetes Care 2019; 42: 406–15.

32 Jia X, Han B, Onengut-Gumuscu S, Chen W-M, Concannon PJ. Imputing Amino Acid Polymorphisms in Human Leukocyte Antigens. PLoS ONE 2013; 8: 64683.

33 Peiris H, Park S, Louis S, et al. Discovering human diabetes-risk gene function with genetics and physiological assays. Nat Commun 2018; 9: 3855.

34 Dos Santos RS, Marroqui L, Velayos T, et al. DEXI, a candidate gene for type 1 diabetes, modulates rat and human pancreatic beta cell inflammation via regulation of the type I IFN/STAT signalling pathway. Diabetologia 2019; 62: 459–72.

35 Pant T, Foda B, Geurts A, Chen Y-G. Lnk/Sh2b3 modulates bioenergetic metabolism of activated CD8 T cells and control the development of Type 1 Diabetes. J Immunol 2023; 210: 77.03.

36 Bottini N, Musumeci L, Alonso A, et al. A functional variant of lymphoid tyrosine phosphatase is associated with type I diabetes. Nat Genet 2004; 36: 337–8.

37 Ämmälä C, Ashcroft FM, Rorsman P. Calcium-independent potentiation of insulin release by cyclic AMP in single β-cells. Nature 1993; 363: 356–8.

38 McGrail C, Chiou J, Elgamal R, et al. Genetic discovery and risk prediction for type 1 diabetes in individuals without high-risk HLA-DR3/DR4 haplotypes. medRxiv 2023; : 2023.11.11.23298405.

39 Redondo MJ, Morgan NG. Heterogeneity and endotypes in type 1 diabetes mellitus. Nat Rev Endocrinol 2023; 19: 542–54.

40 Purcell S, Neale B, Todd-Brown K, et al. PLINK: a tool set for whole-genome association and population-based linkage analyses. Am J Hum Genet 2007; 81: 559–75.

41 McCarthy S, Das S, Kretzschmar W, et al. A reference panel of 64,976 haplotypes for genotype imputation. Nat Genet 2016; 48: 1279–83.

42 Das S, Forer L, Schönherr S, et al. Next-generation genotype imputation service and methods. Nat Genet 2016; 48: 1284–7.

43 SEARCH for Diabetes in Youth: a multicenter study of the prevalence, incidence and classification of diabetes mellitus in youth. Control Clin Trials 2004; 25: 458–71.

44 Danila MI, Laufer VA, Reynolds RJ, et al. Dense Genotyping of Immune-Related Regions Identifies Loci for Rheumatoid Arthritis Risk and Damage in African Americans. Mol Med 2017; 23: 177–87.

45 EPACTS - Genome Analysis Wiki. https://genome.sph.umich.edu/wiki/EPACTS (accessed April 24, 2023).

46 Erlich H, Valdes AM, Noble J, et al. HLA DR-DQ Haplotypes and Genotypes and Type 1 Diabetes Risk: Analysis of the Type 1 Diabetes Genetics Consortium Families. 2008. DOI:10.2337/db07-1331.

47 Noble JA, Valdes AM, Varney MD, et al. HLA Class I and Genetic Susceptibility to Type 1 Diabetes. Diabetes 2010; 59: 2972–9.

48 Varney MD, Valdes AM, Carlson JA, et al. HLA DPA1, DPB1 Alleles and Haplotypes Contribute to the Risk Associated With Type 1 Diabetes: Analysis of the Type 1 Diabetes Genetics Consortium Families. Diabetes 2010; 59: 2055–62.

49 Wakefield J. Bayes factors for genome-wide association studies: comparison with P-values. Genet Epidemiol 2009; 33: 79–86.

50 Zhang K, Hocker JD, Miller M, et al. A single-cell atlas of chromatin accessibility in the human genome. Cell 2021; 184: 5985–6001.e19.

51 Grant CE, Bailey TL, Noble WS. FIMO: scanning for occurrences of a given motif. Bioinformatics 2011; 27: 1017–8.

52 Chen T, Guestrin C. XGBoost: A Scalable Tree Boosting System. In: Proceedings of the 22nd ACM SIGKDD International Conference on Knowledge Discovery and Data Mining. San Francisco California USA: ACM, 2016: 785–94.

53 Elgart M, Lyons G, Romero-Brufau S, et al. Non-linear machine learning models incorporating SNPs and PRS improve polygenic prediction in diverse human populations. Commun Biol 2022; 5: 1–12.

54 Lundberg SM, Lee S-I. A unified approach to interpreting model predictions. In: Proceedings of the 31st International Conference on Neural Information Processing Systems. Red Hook, NY, USA: Curran Associates Inc., 2017: 4768–77.

55 Florkowski CM. Sensitivity, Specificity, Receiver-Operating Characteristic (ROC) Curves and Likelihood Ratios: Communicating the Performance of Diagnostic Tests. Clin Biochem Rev 2008; 29: S83–7.

56 Inshaw JRJ, Cutler AJ, Crouch DJM, Wicker LS, Todd JA. Genetic Variants Predisposing Most Strongly to Type 1 Diabetes Diagnosed Under Age 7 Years Lie Near Candidate Genes That Function in the Immune System and in Pancreatic β-Cells. Diabetes Care 2020; 43: 169–77.

