## Supplementary figures and images for "Genetic association and machine learning improves discovery and prediction of type 1 diabetes"

### Supplemental figures

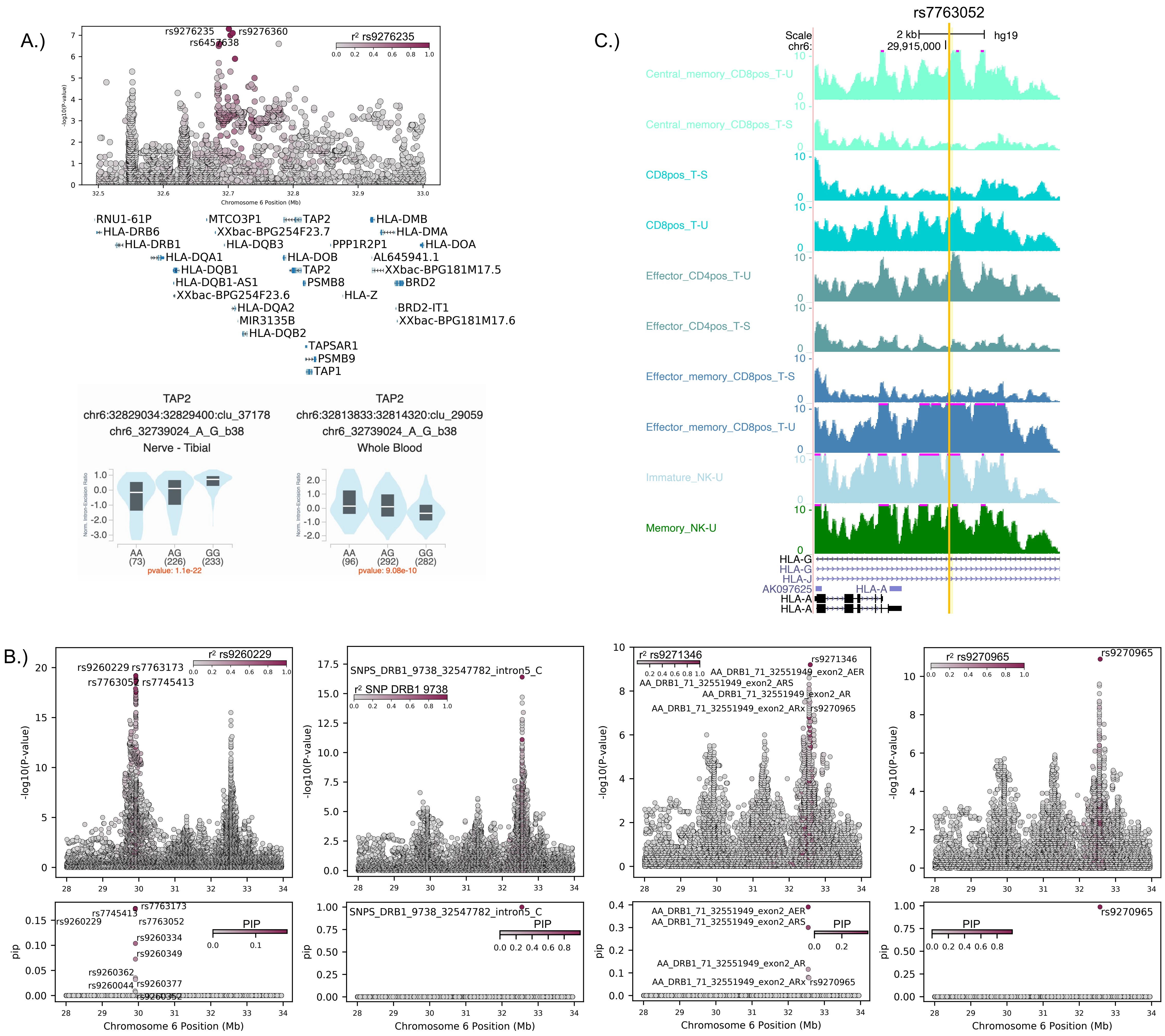

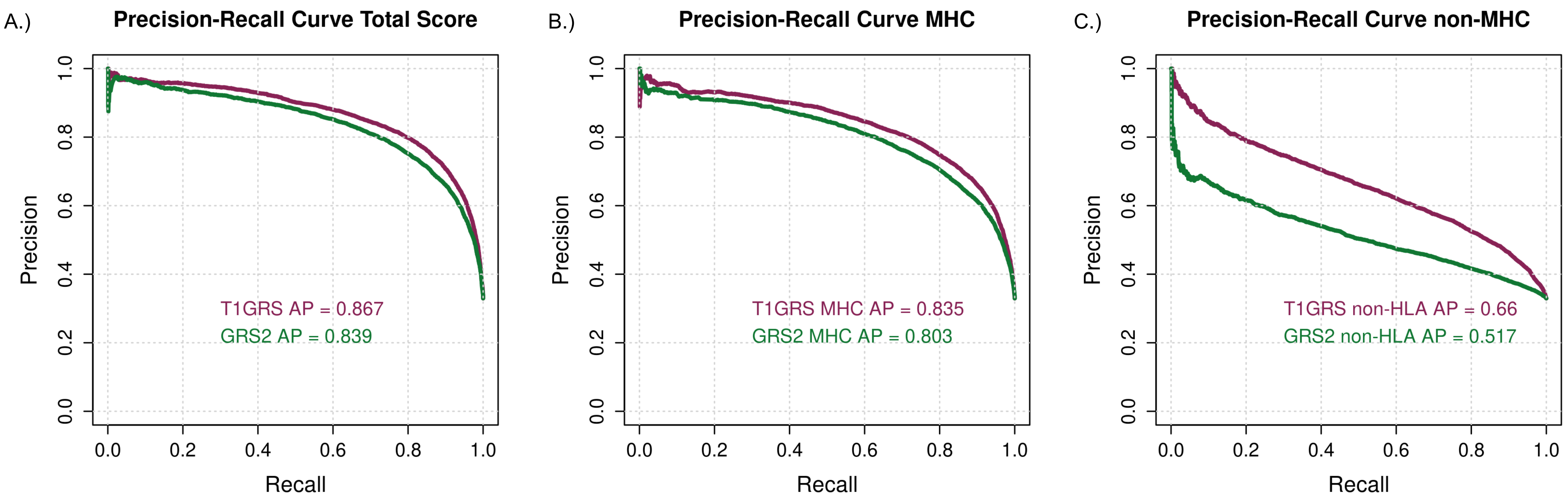

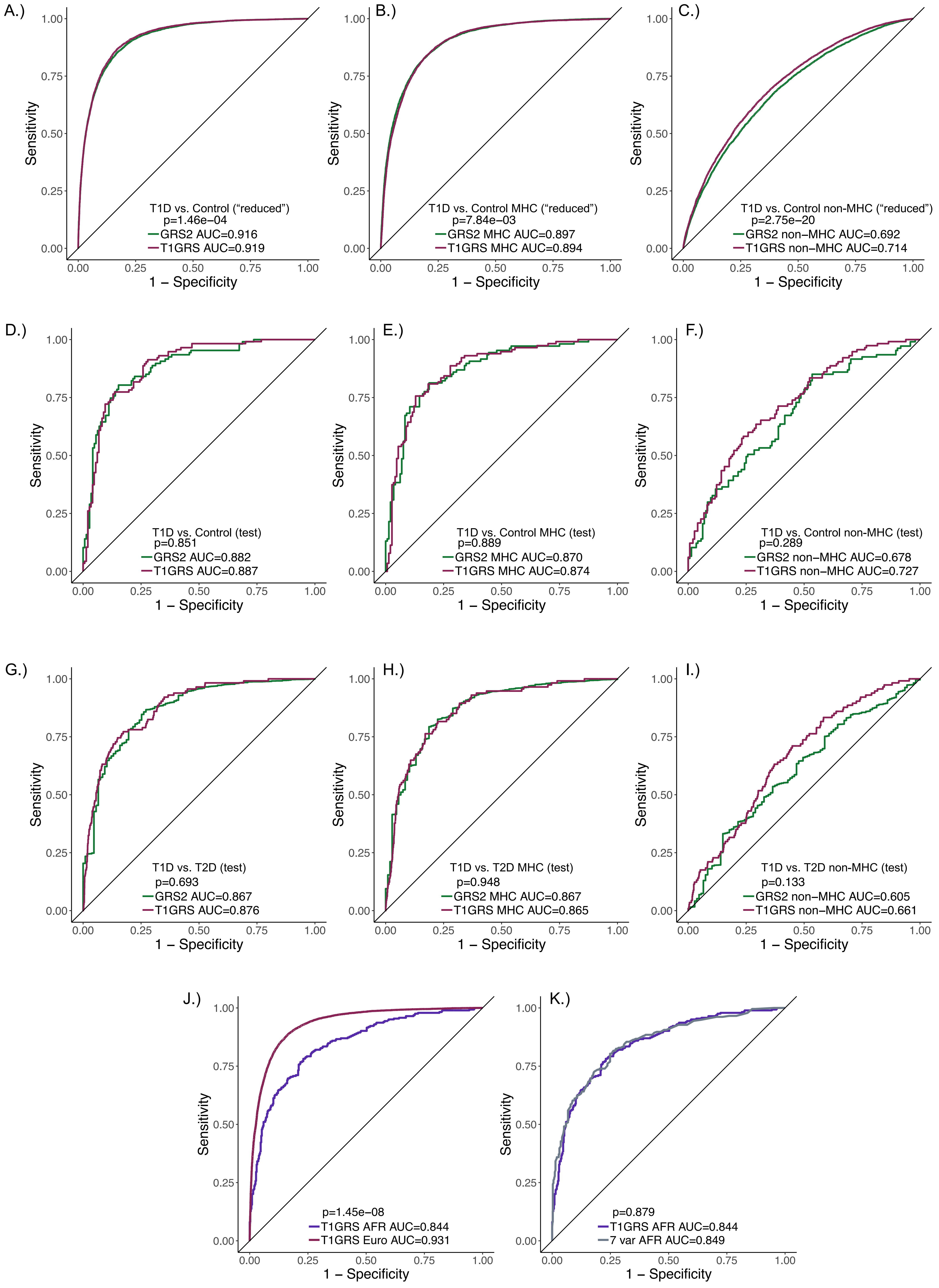


Discovery group: website model no PCs

T1D test (nPOD) vs control

148 Control

115 cases

T1D nPOD T1D (115) vs T2D (1999 WTCCC)

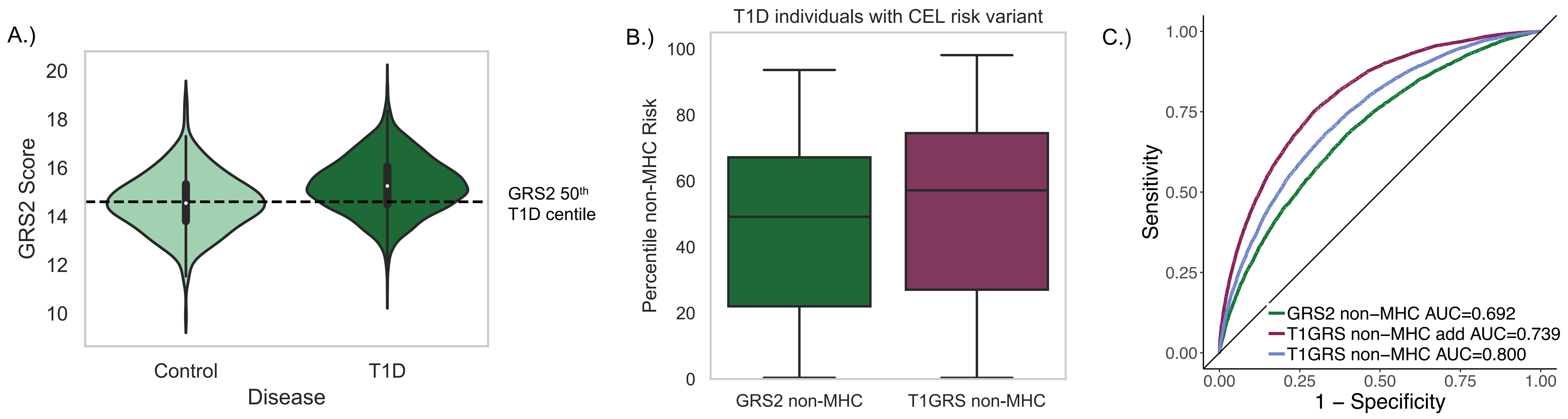

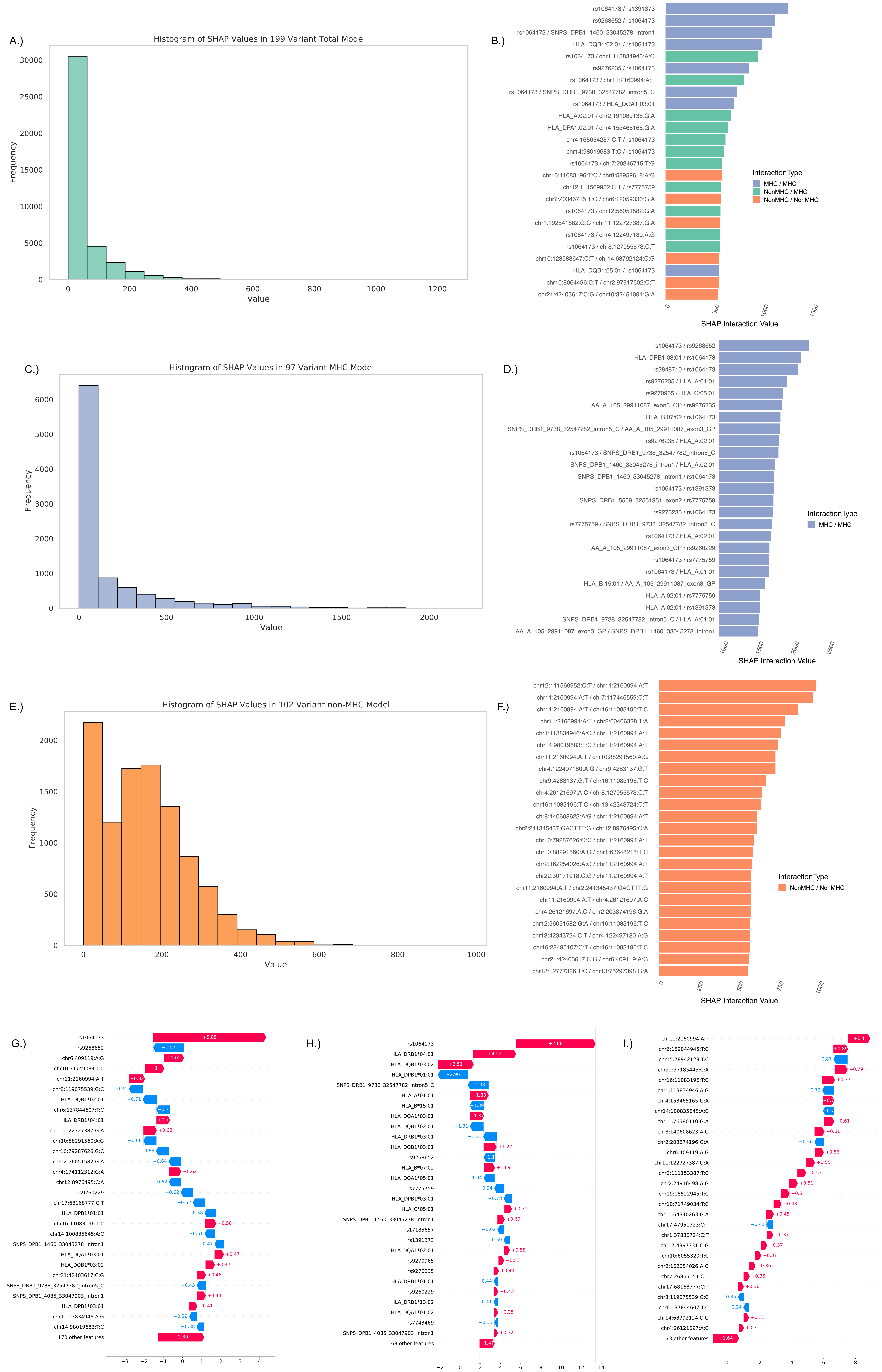

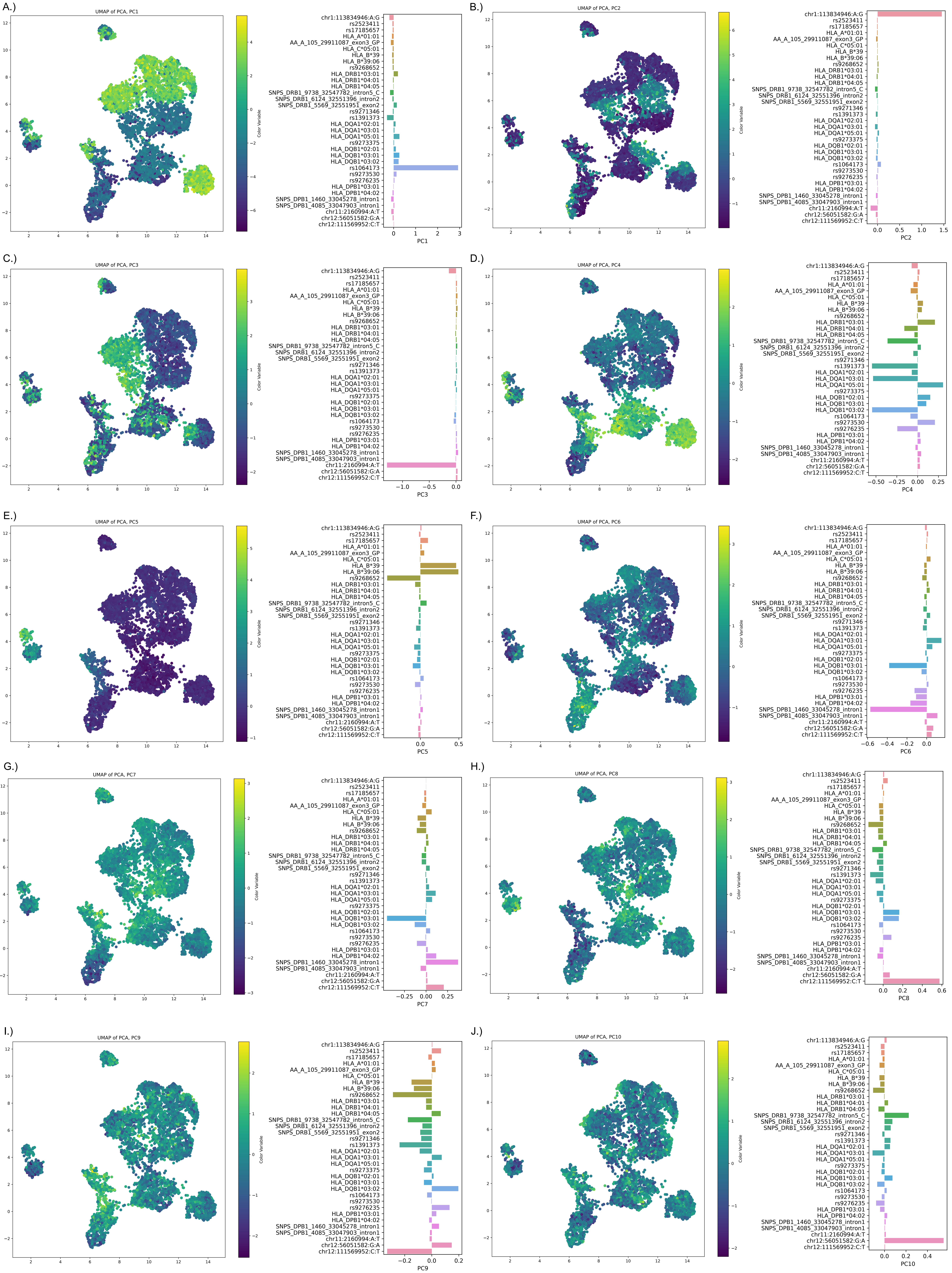
